# Scottish COVID CAncer iMmunity Prevalence (SCCAMP) - a longitudinal study of patients with cancer receiving active anti-cancer treatment during the COVID-19 pandemic

**DOI:** 10.1101/2022.02.22.22271041

**Authors:** Karin Purshouse, John P Thomson, Mahéva Vallet, Lorna Alexander, Isaac Bonisteel, Maree Brennan, David A Cameron, Jonine D Figueroa, Elizabeth Furrie, Pamela Haig, Mattea Heck, Hugh McCaughan, Paul Mitchell, Heather McVicars, Lorraine Primrose, Kate Templeton, Natalie Wilson, Peter S Hall

**Affiliations:** Edinburgh Cancer Centre, NHS Lothian, Crewe Road South, Edinburgh EH4 2XU, UK; Institute of Genetics and Cancer, The University of Edinburgh, Western General Hospital, Crewe Road, Edinburgh EH4 2XU; The University of Edinburgh Medical School, The Chancellor’s Building, Edinburgh BioQuarter, 49 Little France Crescent, Edinburgh, EH16 4SB; Usher Institute, Centre for Population Health Sciences, Old Medical School, Teviot Place, Edinburgh, EH8 9AG; Department of Immunology, Ninewells Hospital and Dundee Medical School, Dundee, DD1 9SY, UK; Clinical Infection Research Group, Regional Infectious Diseases Unit, Western General Hospital, Edinburgh, UK; St John’s Hospital, NHS Lothian, Howden Road West, Howden, Livingston EH54 6PP

## Abstract

**Background:** Cancer and systemic anti-cancer treatment (SACT) have been identified as possible risk factors for infection and related severe illness associated with SARS-CoV-2 virus as a consequence of immune suppression. The Scottish COVID CAncer iMmunity Prevalence (SCCAMP) study aims to characterise the incidence and outcomes of SARS-Cov-2 infection in patients undergoing active anti-cancer treatment during the COVID-19 pandemic and their antibody response following vaccination.

**Patients and Methods:** Eligible patients were those attending secondary care for active anti-cancer treatment for a solid tumour. Blood samples were taken for total SARS-CoV-2 antibody assay (Siemens) at baseline and after 1.5, 3, 6 and 12 months. Data on COVID-19 infection, vaccination, cancer type, treatment and outcome was obtained from routine electronic health records.

**Results:** The study recruited 766 eligible participants between 28th May 2020 and 31st October 2021. The median age was 62.7 years, and 66.5% were female. Most received cytotoxic chemotherapy (79%), with the remaining 14% receiving immunotherapy and 7% receiving another form of anti-cancer therapy (radiotherapy, other systemic anti-cancer treatment). 48 (6.3%) tested positive for SARS-CoV-2 by PCR during the study period. The overall infection rate matched that of the age-matched local general population until May 2021, after which population levels appeared higher. Antibody testing detected additional evidence of infection prior to vaccination, taking the total number to 58 (7.6%). There was no significant difference in SARS-CoV-2 PCR positive test rates based on type of anti-cancer treatment. Mortality proportion was similar between those who died within 90 days of a positive SARS-CoV-2 PCR and those with no positive PCR (10.4% vs 10.6%). Death from all causes was lowest among vaccinated patients, and of the patients who had a positive SARS-CoV-2 PCR at any time, all of those who died during the study period were unvaccinated. Multivariate analysis correcting for age, gender, socioeconomic status, comorbidities and number of previous medications revealed that vaccination was associated with a significantly lower infection rate regardless of treatment with chemotherapy or immunotherapy with hazard ratios of 0.307 (95% CI 0.144-0.6548) or 0.314 (95% CI 0.041-2.367) in vaccinated patients respectively. Where antibody data was available, 96.3% of patients successfully raised SARS-CoV-2 antibodies at a time point after vaccination. This was unaffected by treatment type.

**Conclusion:** SCCAMP provides real-world evidence that patients with cancer undergoing SACT have a high antibody response and protection from SARS-CoV-2 infection following COVID-19 vaccination.

**Highlights:**

- The SCCAMP dataset represents the largest longitudinal study of patients with cancer undergoing anti-cancer treatment during the COVID-19 pandemic
- Rates of infection in the cancer cohort mirrored those of the local age adjusted population
- Vaccination was effective in patients with cancer undergoing active treatment in terms of antibody response and SARS-CoV-2 PCR rates
- Treatment type did not impact the rate of SARS-CoV-2 antibody response

## Introduction

Nearly half a billion people across the world have been infected with SARS-CoV-2^1^. Cancer and systemic anti-cancer treatment (SACT) were identified early as risk factors for infection and related severe illness, particularly given the evidence from previous infection outbreaks^2–4^. A combination of strategies were deployed to protect patients with cancer, including shielding, minimising face to face contact and rationalising treatment regimens^5^. Since then, extensive registry data has highlighted that patients with cancer are at increased risk of mortality from SARS-CoV-2 infection^6,7^. Patients with haematological cancers have been observed to have a higher risk than solid organ cancers of severe SARS-CoV-2 illness, and so evaluating these groups separately is important in understanding the risks posed by SARS-CoV-2 infection^8,9^.

There is concern that immunosuppressive therapy, including SACT, may increase COVID-19 related mortality. Studies in solid organ cancers have shown that male gender, increasing age, presence of comorbidities, performance status and cancer-specific factors such as the extent of tumour burden and lung cancer have been linked with higher COVID-19 mortality risk, but interestingly most studies did not find a relationship between mortality and SACT^5–7^. More recent studies have sought to characterise the immunological response to COVID-19 infection or immunisation in patients with cancer undergoing SACT with varying results. A study capturing data from February to May 2021, including 97 patients with solid-organ cancers and SARS-CoV-2 infection, suggested 89% of patients seroconverted in a cohort where 81% of patients had undergone SACT in the preceding 12 weeks^10^. Further, vaccination was associated with seroconversion rates of 85% after two doses, notably lower than the general population^11,12^. These and other data suggest that patients with solid-organ cancers do broadly develop an immune response to SARS-CoV-2, although it may be slightly reduced^10,11,13^. Other studies estimate an even lower or delayed immune response in this group of patients, although studies often include patients with haematological cancers, have early antibody testing strategies and may lack long-term follow-up^13–16^. Real-world, longitudinal data is still needed. Overall, given the importance of maintaining anti-cancer care in an age where SARS-CoV-2 is endemic, it is important to understand the immune response to both SARS-CoV-2 infection and COVID-19 vaccination in patients being treated for cancer.

The Scottish COVID CAncer iMmunity Prevalence (SCCAMP) study aims to comprehensively assess COVID-19 infection as proven by standard-care RT-PCR and to use a SARS-CoV-2 antibody test, alongside linked data from electronic health records, to assess response to infection and vaccination in patients undergoing active cancer treatment.

In this preliminary report we describe outcomes in a cohort of patients receiving active cancer treatment during the COVID-19 pandemic between May 2020 and October 2021 who have contributed serial blood samples for antibody testing when attending for treatment.

## Methods

### Study design

The SCCAMP study protocol is available on https://cancer-data.ecrc.ed.ac.uk/projects/sccamp/sccamp-information-for-professionals/. Patients were eligible if they were over the age of 18 with a confirmed diagnosis of solid organ cancer, defined as cancer or metastasis in situ, and/or receiving cancer treatment (surgery, radiotherapy, hormone therapy, chemotherapy, targeted therapy, immunotherapy) in the last 12 months, and attending for outpatient Cancer Centre Care. Patients were not eligible if they had a concurrent haematological malignancy due to the different clinical profile of this cohort. Consent was provided when attending for anti-cancer treatment (ACT), primarily SACT, at the Edinburgh Cancer Centre (ECC) either at the Western General Hospital (WGH), Edinburgh or St John’s Hospital (SJH), Livingston (NHS Lothian NRS BioResource, BioBank SR1418, NHS Research Ethics Committee (REC): 20/ES/0061 and SCCAMP, NHS Research Ethics Committee (REC) REC: 20/SS/0109). Blood samples were taken for antibody testing at consent up to a maximum of five collections up to 1 year from consent (approx. +42 days, +84 days, +6 months, +1 year), when returning for further routine out-patient care.

### Data collation

Clinical information was obtained through data linkage from routine Electronic Patient Records including prescribing systems (ChemoCare™ - https://www.scan.scot.nhs.uk/projects/chemocare/), PCR/vaccine data was obtained from Public Health Scotland (https://www.publichealthscotland.scot/), and comorbidity data was obtained from SMR01 (General/Acute Inpatient and Day Case - https://www.ndc.scot.nhs.uk/Data-Dictionary/SMR-Datasets/SMR01-General-Acute-Inpatient-and-Day-Case/) and Prescribing Information System (PIS - https://www.ndc.scot.nhs.uk/National-Datasets/data.asp?SubID=9). All patient data was compared to their recruitment date which coincided with their first blood sample date (referred to as the patient’s “baseline date”). Canc_er type at recruitment was extracted and stratified into one of 8 groups based on the most dominant cancer types seen in the study (see supplemental methods). Socioeconomic status was calculated from residential postcodes at recruitment which were cross referenced to Scottish Index of Multiple Deprivation (SMID) scores, binned into quintiles (1 = low, 5 = high socioeconomic status). Quan-Charlson indices (QCIs) were calculated using the weightings of Quan et. al.^17^ but excluding cancer as a comorbidity. This was used to define 5-year comorbidities occurring prior to the patients’ Scottish Incidence date, as recorded in the cancer registry (https://www.ndc.scot.nhs.uk/National-Datasets/data.asp?ID=5&SubID=8). Total prescribed medicines within 1 year prior to consent were also extracted. Treatment regimens were hierarchically classified into one of 3 classes (chemotherapy > immunotherapy > other) for the duration of the study including 6 months prior to recruitment. Patients receiving more than one therapy were classified by their hierarchy. Other treatments include therapies such as radiotherapy, hormone treatment and small molecule treatment (see supplementary table 1 for full drug list).

All data was up to date as of 31st October 2021, at which point data was censored.

### COVID-19 data

COVID-19 positive cases were defined as cases with a supporting positive PCR test. Publicly available population COVID-19 data was accessed from Public Health Scotland (PHS) data sources at (www.opendata.nhs.scot/dataset/covid-19-in-scotland) and monthly incident rates and cumulative total calculated for the combined local authorities in which the two hospital sites reside (the City of Edinburgh and West Lothian). Population COVID-19 infection rates were adjusted to per 1000 population values based on census data accessed from the National Records of Scotland (NRS - https://www.ndc.scot.nhs.uk/National-Datasets/data.asp?ID=3&SubID=13), or for cancer patients the total study size. As the ages of the patients in our cohort are all >25 years old, population COVID-19 data was age corrected to remove individuals under the age of 25 (see supplemental methods). Vaccination data within the cancer cohort was provided by PHS.

### COVID-19 antibody testing and analysis

Serum samples were tested via the validated Siemens Total (IgG/M and IgA) SARS-CoV-2 antibody assay at Ninewells Hospital, NHS Tayside with thresholds for antibody applied as previously defined^18,19^. For full methods on antibody class stratification and analysis see supplementary methods.

To calculate antibody responses in total and split by treatment type, we considered cases with either a positive antibody result in the first collection after the date of first vaccination or a positive or negative antibody result >14 days after the date of second vaccination. To calculate COVID-19 prevalence between fully and partial/non vaccinated states, we considered all cases with at least 1 available antibody test (pre-vaccination only) and/or a positive PCR test. To discriminate between COVID-19 infection and vaccination induced seroconversion, vaccination status was considered at the time of either a positive PCR or seroconverted antibody test result.

### Data visualisation and statistical analysis

All analysis was carried out using base R version 4.0.5. For all univariate and multivariate analysis, COVID-19 positive cases were only considered if they occurred after the date of most recent cancer treatment (43/48 cases). Univariate and multivariate analysis was carried out using the Survival package 3.2-13 in order to compute the Cox proportional hazards regression models.

A multivariate model investigating the risk of catching COVID-19 during the study was defined as the length of time free from infection with respect to recruitment (day) with patients without a COVID-19 positive PCR censored and the following variable binary groupings applied: Age > 60, gender = female, high socioeconomic score = SMID quintiles 4 & 5, High medication comorbidity > 5 prescribed medications in 1 year prior to recruitment, QCI score > 0 in 5 years prior to recruitment, cancer treatment class (chemotherapy, immunotherapy and other) as a binary yes/no events, vaccinated = 2 or more doses. P values were calculated by performing a two-proportion Z-test.

## Results

### Patient demographics

767 patients attending for ACT consented between 28th May 2020 and 28th October 2021, of whom 766 were included for analysis (See Consort diagram). Patients were recruited across two sites within Edinburgh and West Lothian (612, 79.9%, at the Western General Hospital in Edinburgh and 154, 20.1%, at St John’s Hospital in Livingstone) with a median follow up of 405 days (min 3, max 521) [Figures 1A & 1B, Table 1].

**Table 1:** summary of patient data in the SCCAMP study

|  |  | N | % of total cohort |
| --- | --- | --- | --- |
| Cases | Total | 766 |  |
| Follow up period | Median days | 405 | range 3-521 |
| Centre | Western General Hospital Edinburgh | 612 | 79,9 |
|  | St Johns Hospital, Livingstone | 154 | 20,1 |
| Age at recruitment | Median years | 62,8 | range 26-87.8 |
| Gender | Female | 510 | 66,6 |
|  | Male | 256 | 33,4 |
| Cancer type | Breast Cancer | 289 | 37,7 |
|  | Lung and chest | 98 | 12,8 |
|  | Gynae | 94 | 12,3 |
|  | Lower GI | 92 | 12,0 |
|  | Upper GI | 63 | 8,2 |
|  | Urilogical | 51 | 6,7 |
|  | Skin | 38 | 5,0 |
|  | Other | 41 | 5,4 |
| Socioeconomic status (SIMD Quintiles) | 1 | 82 | 10,7 |
|  | 2 | 153 | 20,0 |
|  | 3 | 130 | 17,0 |
|  | 4 | 129 | 16,8 |
|  | 5 | 272 | 35,5 |
| Comorbidity: Quan-Charlson score ≤ 5yr | 0 | 690 | 90,1 |
|  | 1 | 51 | 6,7 |
|  | 2 | 20 | 2,6 |
|  | ≥3 | 5 | 0,7 |
| Comorbidity: Prescribed meds ≤ 1 yr | Median prescribed meds | 5 | range 0-28 |
| Vaccination status at end of study or at death | ≤1 | 155 | 20,2 |
|  | ≥2 | 611 | 79,8 |
| PCR confirmed COVID-19 | Yes [<6M prior to treatment] | 6 | 0,8 |
|  | Yes (during treatment) | 42 | 5,5 |
|  | No | 718 | 93,7 |
| Treatment intent | Palliative | 441 | 57,6 |
|  | Curative | 325 | 42,4 |
| Treated with chemotherapy | Yes | 603 | 78,7 |
| Treated with immunotherapy; no chemotherapy | Yes | 107 | 14,0 |
| Other treatment; no immunotherapy, no chemotherapy | Yes | 56 | 7,3 |
| Death details | Died within 28d of PCR confirmed COVID-19 | 2 | 0.3 [4.2% COVID-19 positive] |
|  | Died within 90d of PCR confirmed COVID-19 | 5 | 0.7 [10.4% COVID-19 positive] |
|  | Died < 2 years after PCR confirmed COVID-19 | 9 | 1.9 [18.7% COVID-19 positive] |
|  | Died, no PCR confirmed COVID-19 | 158 | 20,6 |
|  | Alive at end of study | 592 | 77,3 |

**Figure 1.**
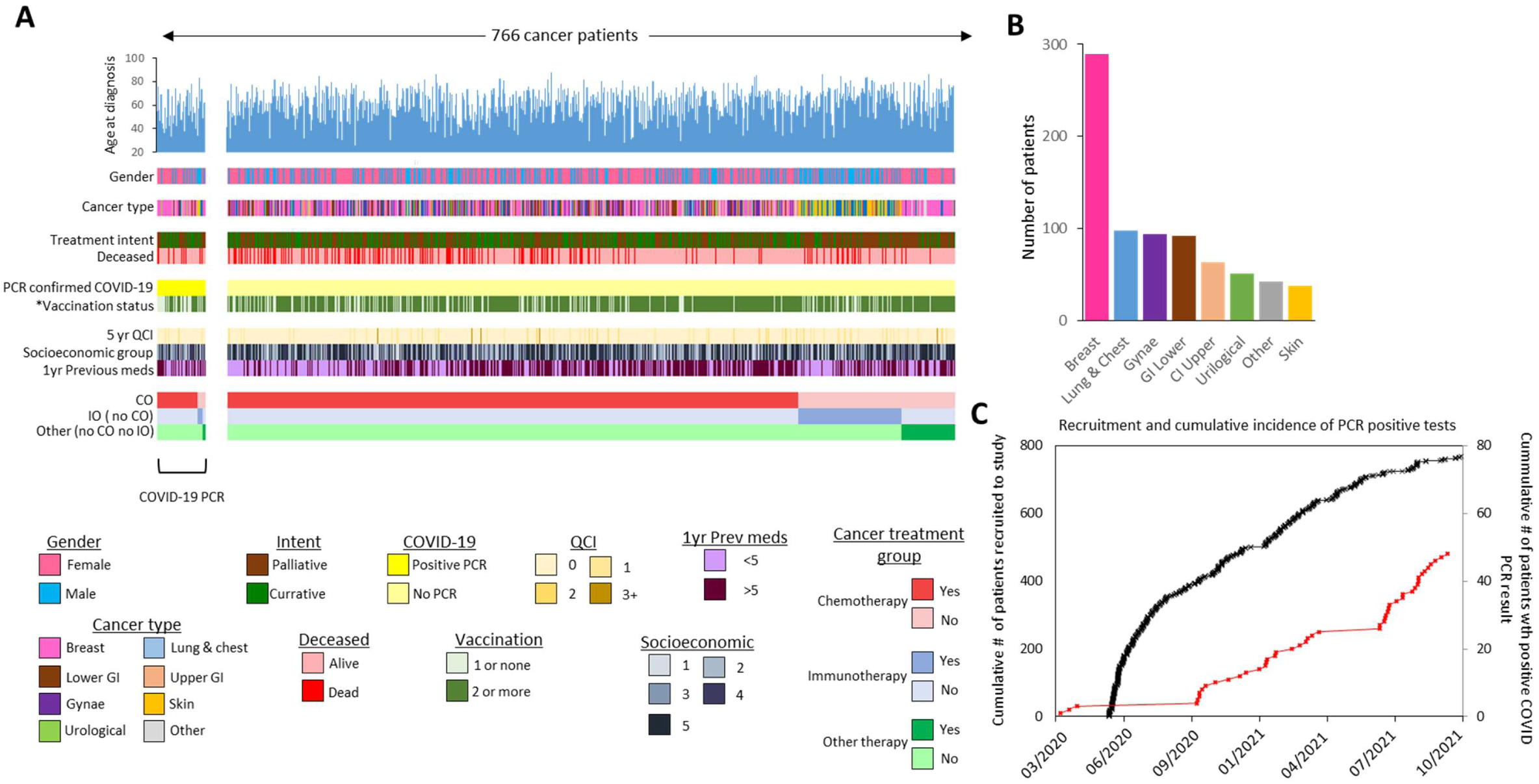
**A**. Graphical summary of available data in the SCCAMP study across the 767 patients. Patients are stratified by their COVID-19 PCR status (yellow) and then ranked by their treatment type. The Colour key for each data type is shown below. * vaccination status is defined as status at time of COVID-19 positive PCR or end of study if not positive.**B**. Plot of patient numbers split by cancer type. **C**. Plot of patient recruitment throughout the study (black line) with confirmed cases of COVID-19 overlaid (red line).

The median age across the cohort was 62.8 years (min 26, max 87.8) with 510 females (66.6%) and 256 males (33.4%) [Supplemental figure S2]. The cohort was composed of cancer patients with solid tumours falling into one of 8 classifications: cancers of the breast (n = 289), lung and chest (n = 98), gynaecological (n = 94), lower gastrointestinal (n = 92) and upper gastrointestinal (n = 63) [Figures 1A & 1B, Table 1 & Supplementary figure S3]. Number of patients in each socioeconomic quintile as defined by SIMD quintile scores (Q1 low socioeconomic - Q5 high socioeconomic) were Q1: 82, Q2: 153, Q3: 130, Q4: 129 and Q5: 272 respectively [Figure 1A, Table 1 & Supplemental figure S4]. Five year comorbidity as described by QCI score defined the vast majority of patients (n = 690; 90.1%) as being without any associated comorbidity. The remaining 76 patients had QCI scores ranging from 1 (n = 51), 2 (n = 20) or >2 (n = 5). QCI scores associated with previous medication (1 year pre-recruitment) was also investigated across the patients revealing a median of 5 previous prescribed medications (range 0-28).

Overall 325/766 patients (42.4%) were being treated with curative intent. Across the duration of the study, 603/766 (78.7%) were classified as receiving cytotoxic therapy, 107/766 classified as receiving immunotherapy (in the absence of cytotoxic therapy; 14%) and 56/766 (7%) classified as receiving another treatment in the absence of cytotoxic and immunotherapeutic intervention [Figure 1A & Table 1]. 497/766 (64.9%) patients received more than one therapeutic intervention type [Supplemental Figure S5].

### COVID19 infection rates within the cancer cohort

Over the period of the study, 48/766 cancer patients (6.3%) had a recorded positive COVID-19 PCR test. [Figure 1B & 2A]. 5 patients tested positive for COVID-19 prior to the start of their cancer treatment however these individuals went on to receive treatment within 6 months of infection. Excluding these 5 cases, median time from first cancer treatment to COVID-19 infection was 230 days (min 2 days, max 638 days). Ten of the 48 cases remained positive in at least one follow-up PCR test (median follow up 7 days, low 1, high 21) [supplemental figure S6]. No cases of reinfection with COVID-19 were seen. Cases were found across 7 of the 8 cancer type groups, although the proportion of cases in each category were not randomly distributed (Chi squared test = 5.3E-05), with more cases in those with breast cancer [supplemental figure S3].

In comparing COVID rates within the cancer patients against age adjusted NRS population data over the same geographic area, the cumulative incidence as well as the incidence rate of cases was broadly the same between our cancer patient cohort and the general population until approximately May 2021, when the proportion started to increase in the general population relative to our cohort [Figures 2B & 2C]. The age adjusted cumulative incidence in May 2021 was 3.4 and 2.9 per 1000 for the local general population and the SCCAMP cancer cohort respectively and by the end of the study 9.6 per 1000 for the local population and 5.9 per 1000 in the cancer cohort [Figures 2B & 2C].

**Figure 2.**
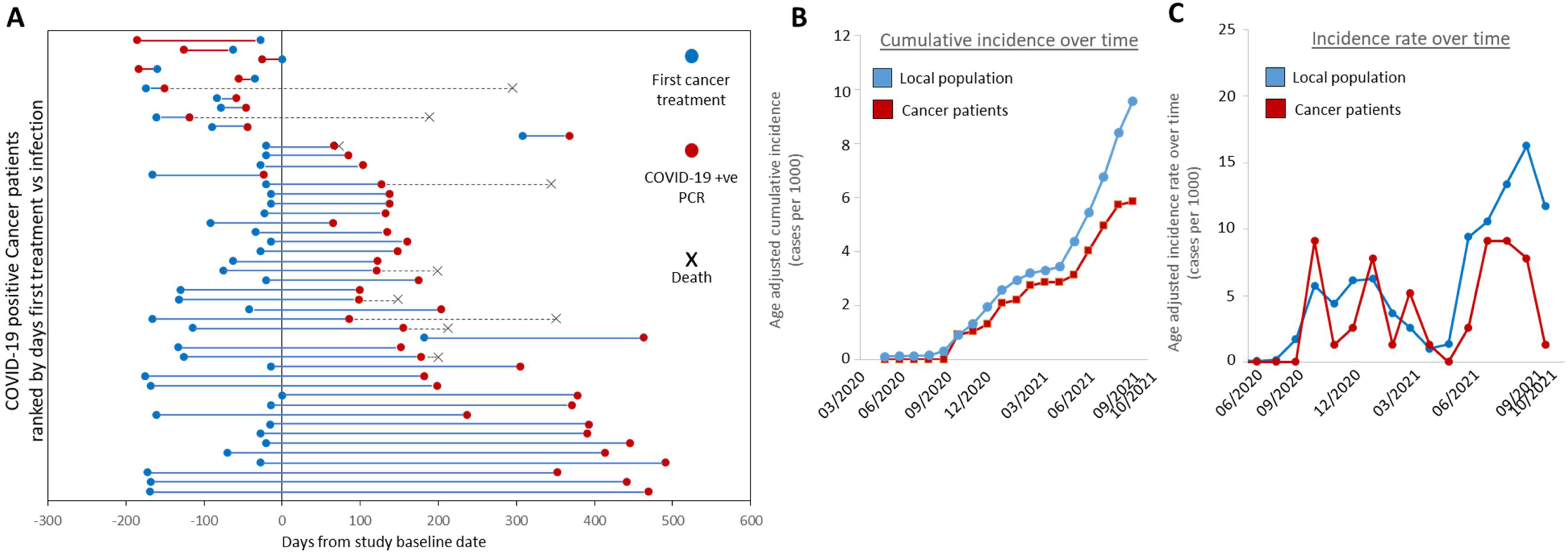
**A**. Plot of time between first cancer treatment (blue dots), COVID-19 infection (red dots) and death (black cross) across the 48 COVID-19 positive cancer patients in the study ranked by time between treatment and infection. Red bars denote cases where infection occurred prior to first treatment, blue bars where infection occurs after first treatment. Plots display time with respect to date of recruitment into the study. **B**. Age adjusted COVID-19 cumulative incidence over time and **C** incidence rate over time plots for SCCAMP patients (red) and the local general population (blue). Values are presented as PCR confirmed cases per 1000 per month.

### Mortality and association of cancer treatment with COVID-19 infection

Across the study period, 2/48 patients (4.16%) died within 28 days of a positive PCR result, one of which had a urological cancer, the other non-small cell lung cancer (NSCLC) (Supplemental figure S7). Expansion of this period to include all deaths within 90 days resulted in a total of 5 deaths (10.4%) and expansion to include all deaths recorded across the study period after a COVID positive test resulted in 9 total deaths (18.7%, median survival days from recruitment 200). 8/9 (88.9%) deaths registered at any time after a COVID-19 infection were being treated with palliative intent. By contrast 158 cancer patients who did not report a COVID-19 positive PCR result died over the entirety of the study period (20.6% total cohort. median survival days from recruitment 192), 86.8% of whom were being treated with palliative intent [Table 1].

Median time from treatment initiation to COVID-19 infection was 196 days across these 9 patients (min 23 days, max 304 days) which compares to 246 days for patients who were alive at the end of the study (excluding cases where COVID-19 was contracted prior to treatment initiation) although this did not reach significance (2 tailed t-test P-value 0.079) [Figure 2A & Supplemental figure S8].

COVID positive cancer patients tended to be younger than non-positive cases [supplemental figure S2] although there was no significant difference between the ages of all cancer patients who died during the study to those who died either at any time after COVID-19 infection, or within 28 days [supplemental figure S2]. Interestingly, patients who had experienced a COVID-19 infection who died within the first half of the study exhibited significantly shorter times between infection to death than those in the second half of the study (First 9 months: n=4, May 2020 - Jan 2021, median days from infection to death = 40; second 9 months: n=5, Feb 2021 - Oct 2021, median days from infection to death = 242; two tailed T-test P-value =0.023).

There was no significant difference in SARS-CoV-2 infection rates depending on treatment type received. Excluding cases where COVID-19 was contracted prior to treatment initiation (n=5), positive COVID-19 PCR rates between patients in the chemotherapy treatment group were 6% (36/599), 3.8% in the immunotherapy group (4/106) and 5.4% (3/56) in those who received other treatments [Table 2]. In adjusted models for COVID-19-free days there was no significant difference in positive COVID-19 PCR rates by treatment group (Chemotherapy: Hazard Ratio (HR) 1.41 (95% CI 0.63-3.18); immunotherapy: 0.62 (95% CI 0.22-1.74); other treatments: HR 0.94 (95% CI 0.29-3.02); chemotherapy without immunotherapy: HR 1.71 (95% CI 0.80-3.72)).

**Table 2:** COVID-19 PCR rates per treatment type

|  | Chemotherapy | Immunotherapy | Other therapy | Total |
| --- | --- | --- | --- | --- |
| COVID-19 positive | 36 | 4 | 3 | 43 |
| COVID-19 negative | 563 | 102 | 53 | 718 |
| % COVID-19 positive | 6.0% | 3.8% | 5.4% | 5.7% |

Patients who had a positive COVID-19 PCR were younger and less likely to be double vaccinated (>60yr age: 58.3% no COVID-19 vs 45.9% COVID-19 +ve, 12.3% decrease in COVID-19 infected patients; (Double vaccination: 79.1% no COVID-19 vs 59.5% COVID-19+ve, 19.7% decrease in COVID-19 infected patients). There was a less than 10% difference between those who tested positive and the remaining cohort in the following factors: QCI scores, numbers of previous medications, socioeconomic status scores, cancer treatment class and gender (Table 3).

**Table 3:**
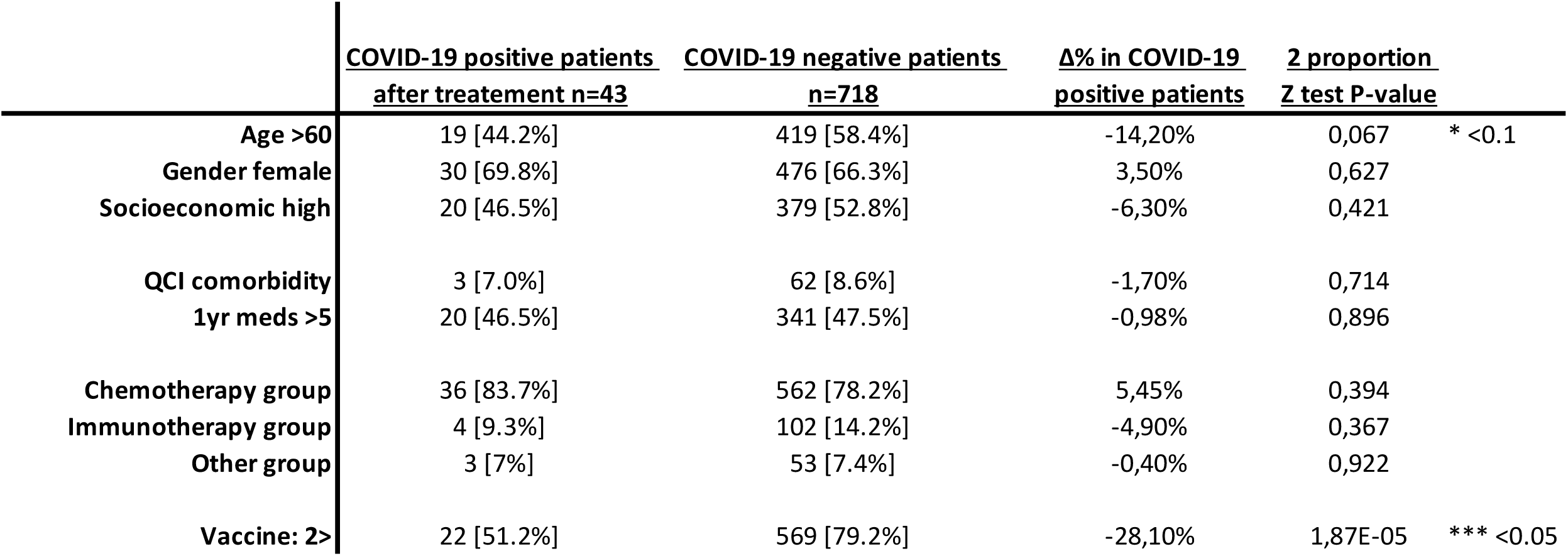
Patients characteristics

Multivariate analysis adjusting for age, gender, QCI scores, previous medications and socioeconomic factors reveals that vaccination reduced the risk of having a positive COVID-19 PCR test across the entire patient cohort (HR 0.26 (95% CI 0.14-0.48)). More specifically, this is not dependent on treatment by either chemotherapy only or immunotherapy only; with hazard ratios of 0.21 (95% CI 0.10-0.41) or 0.314 (95% CI 0.041-2.37) respectively.

### COVID19 vaccination in the cancer cohort

The COVID-19 vaccination programme began in Scotland on 8th December 2020, with patients with cancer among those prioritised. 730/766 patients from our cohort were alive when the vaccination programme began (95.3%). By the date of data censoring, 155 patients received less than 2 vaccinations (20.2% of the cohort). Among 117 unvaccinated patients, 59% died; 32% died before the programme began (n=37) and 27% died within 6 months of first vaccine (n = 32). By contrast 246 had received 2 vaccine doses (32.1%) and 365 had received 2 doses plus a booster (47.7%) [Figures 3A, 3B & Figure 4A]. As such 79.8% of the cancer cohort received at least 2 vaccine doses over this period of time. This compares to vaccination rates of 71.5% for 2 vaccines and 13.2% for <2 vaccines across the national population of Scotland over the same time period^20^. The majority of patients in the study received Astrazeneca vaccines either as their first (88.2%) or second (87.6%) vaccine with a minority receiving Pfizer as their first (11.7%) or second (12.1%) dose. By contrast the majority of booster vaccines were either Pfizer (65.2%) or Moderna (34.5%) [Figure 3A].

**Figure 3.**
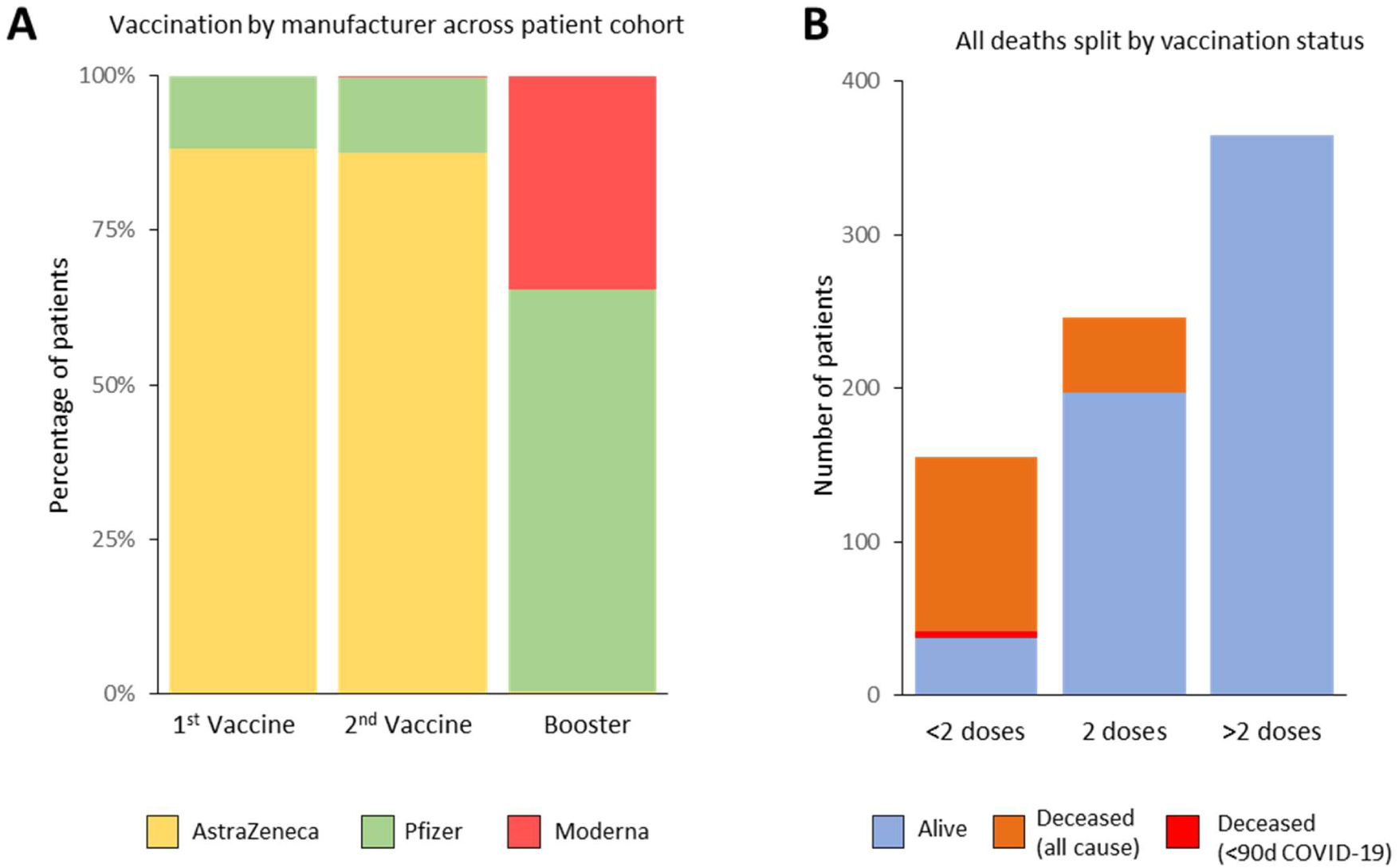
**A**. Stacked bar plot showing the percentages of the total cohort split by the manufacturer of the vaccine received at 1st dose, 2nd dose or booster. **B**. Plot of total vaccine doses per patient across the study with mortality data overlaid as a stacked plot. Alive = blue, deceased = orange, deceased within 90 days of confirmed Covid-19 infection = red.

**Figure 4.**
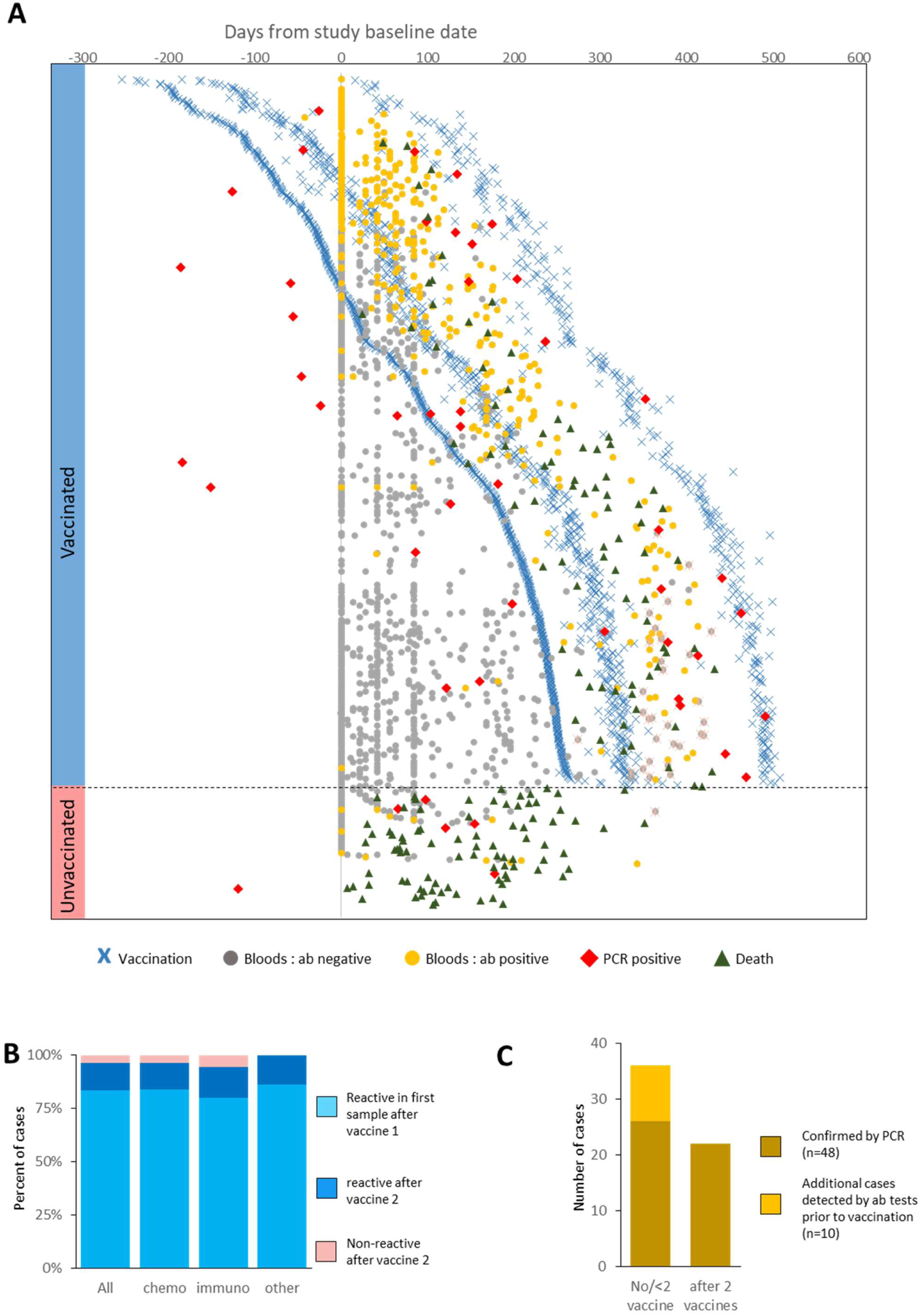
**A**. Plot of timings of vaccinations, antibody data points and PCR test results for the 766 patients with respect to baseline dates. Patients are arranged in order of their first vaccination date. Vaccinations are denoted by crosses (light blue = 1st, dark blue = 2nd, green = 3rd). Dots denote antibody test collection dates w.r.t. Baseline data. Grey dots = antibody non-reactive result, yellow dots = antibody reactive result, red dot = date of PCR positive test. Blue bar on the left denotes patients who had received at least 1 vaccine dose, the pink bar at the bottom left denotes unvaccinated patients. Dashed line separates these two groups. **B**. Stacked bar plot of percentage of patients reporting antibody reactivity split by response type. Data is split for all available patient data as well as by treatment class. Light blue= Reactive in first sample after vaccine 1, dark blue= reactive after vaccine 2, pink = Non-reactive after vaccine 2. Data shows samples for which there is either a reactive antibody result in the first sample after vaccine 1 and/or antibody data available >14 days after second vaccination. **C**. Plot of number of COVID-19 cases confirmed by PCR test (gold) or suspected infection through antibody reactivity prior to vaccination, split by patient vaccination status.

Proportions of deaths from all causes differed across these three groups with a death recorded for 118/155 non-fully vaccinated patients (76.1%), 49/256 double dosed patients (20%) and no deaths recorded for triple dosed patients. Notably, the only cohort in which COVID-19 related deaths were reported (classified here as a death recorded <90 days after positive PCR) were unvaccinated individuals (n=5), two of which were within 28 days [Figure 3B].

### Antibody positive over time and cumulative incidence

Next we looked at COVID-19 seroconversion in our cohort to assess antibody response to vaccination and undetected COVID-19 infection events. At the time of analysis, antibody data for at least 1 time point was available for 591 patients (77.1% of cohort), with a total of 1418 samples collected longitudinally at baseline and then at 1.5, 3, 6 and 12 month follow up periods. Across these 591 patients, 348 (45.4%) contained data which overlapped with time points at least 14 days after the date of first vaccination [Figure 4A]. In total we collected a median of 2 (min 1, max 5) antibody samples per patient over a median span of 95 days (min 14 days, max 429 days) [supplemental figure S9]. At least 1 reactive antibody result was noted in 304/589 patients (51.6%) which was restricted to 285/347 patients (82.1%) for which antibody data was at least 14 days after vaccination 1.

We defined the cohort of patients in whom we could assess seroconversion post vaccination as being patients with at least 1 reactive antibody result after vaccination 1 or at least 1 antibody test >14d after 2nd vaccination (n = 297). We report that 248/297 (83.5%) patients display a reactive result in the first blood sample after vaccination 1 [Figure 4B]. 38/297 cases display an initial non-reactive result >14 days after their first vaccination prior to becoming reactive at a later stage (12.8%) (7/38 pre-second vaccination, 31/38 post second vaccination), with only 11 patients returning no positive results >14 days post second vaccination [supplemental figure S10, S11]. We therefore observe an antibody response rate to vaccination of 96.3%. No differences were observed when stratifying these response classes by treatment type (Figure 4C), nor by vaccination manufacturer [Supplemental figure 12].

Across all samples we observe only 4 cases where seroconversion was lost at a later date; 3 of whom had received 2 doses of the vaccine at time of reversion (median time to reversion at 42 days after a previously positive result) and one case who reverted prior to their first vaccination (asymptomatic infection) [Supplemental figure S13].

Finally, through the analysis of antibody data we note a number of additional COVID-19 infections, prior to any vaccination, which were not detected by PCR tests (n=10) [Figure 4A]. Combining these additional pre-vaccination cases with PCR confirmed COVID-19 cases results in 36 positive cases in patients prior to a 2nd vaccination (26 PCR positive, 10 additional pre vaccination antibody reactive) compared to 22 PCR positive cases in patients after their 2nd vaccination [Figure 4C].

## Discussion

We describe the findings of the SCCAMP study which seeks to characterise the pattern of SARS-CoV-2 infection and subsequent immune response in a cohort of patients with cancer undergoing anti-cancer treatment between May 2020 and October 2021.

Studies have previously highlighted a higher mortality risk for patients with cancer, and proposed that certain groups of patients are at higher risk of severe SARS-CoV-2 infection, such as those with advanced disease or lung cancer^5–7,21,22^. Many early studies could be influenced by a highly diverse definition of cancer and, importantly, changes in anti-cancer treatment policies during the pandemic^5^. More recent studies have evaluated the immune response of patients with cancer following SARS-CoV-2 infection and vaccination, with a wide variation in proposed responses owing to differences in study populations^10,11,13–15^. In contrast with much of the UK, the South East Scotland Cancer Network (SCAN), which comprises both hospitals reported here, had largely normalised SACT attendance rates by June/July 2020 (vs −31.2% in England and Northern Ireland)^23,24^. SCCAMP therefore offers an opportunity to understand trends in SARS-CoV-2 infection and immunity in a cohort of cancer patients similar to that of a pre-COVID-19 era, and during a period of multiple waves of SARS-CoV-2 variants. We report trends in COVID-19 incidence in this population, in addition to immunity patterns, in patients who are deemed well enough to undergo anti-cancer treatment, are outpatients and were asymptomatic at the time of sampling. This is important given the likelihood of ongoing waves of SARS-CoV-2, and the need to continue anti-cancer treatment to avoid the risk of cancer mortality exceeding that of SARS-CoV-2.

Our data demonstrate that an actively treated cohort of patients with cancer had a similar incidence of COVID-19 infection to the regional population. The small number of cases in our cohort makes it difficult to make inferences about overall risk, but we did not observe a higher risk depending on treatment type, in keeping with other studies^6,7^. The mortality proportion between those who ever and never had a positive COVID-19 PCR test were broadly similar, although mortality from SARS-CoV-2 infection was notably lower than in previous observational studies^5–7^. This likely reflects the relative fitness of this cohort, given that all patients recruited here were asymptomatic on the days they gave samples, were outpatients, had few comorbidities and were deemed fit enough for ACT. In patients who tested positive for SARS-CoV-2 and died during the follow-up period, the time between SARS-Cov-2 infection and death was shorter in those diagnosed during the first half of the study. This may be reflective of vaccination, advances in the management of SARS-Cov-2 illness or changes in the patient cohort, although numbers are small. We also had relatively few patients with lung cancer, a group considered to harbour higher risk from SARS-CoV-2 infection, which may also have influenced this result.

After approximately May 2021, the rates of SARS-CoV-2 infection in the regional population were higher than in the SCCAMP cohort, suggesting a protective effect from vaccination given that patients with cancer were a priority group. Despite our small rate of SARS-CoV-2 infection, we still observed a significant reduction in the number of positive COVID-19 PCR results in patients who had received 2 or more doses of any COVID-19 vaccination. Taken together, these data highlight that patients undergoing active treatment for cancer gain significant protection from SARS-CoV-2 infection by receiving a COVID-19 vaccination.

We observed a high rate of vaccination in our cohort, with nearly 50% of patients having received a booster COVID-19 vaccination at the time of censoring (Oct 31st) in comparison with the Scottish rate (13.2% - scot.gov), and an overall higher proportion of people having received at least two doses than the general population at the same time. The proportion of patients (20.3%) that had received no or a single vaccine is likely reflective of patients who died of their cancer or related causes prior to receiving both doses. Our data suggest that most patients with cancer who are receiving ACT are both being reached and are engaged with COVID-19 vaccination.

We observed that 96.3% of patients in our cohort who were vaccinated had seroconverted when considering any positive result post first vaccination, and all results >14 days post second vaccination as the denominator. This is higher than reported in some studies^11,14,15^. Importantly, there was no difference in response between patients receiving chemotherapy, immunotherapy or other treatments. Others have reported that patients with cancer may have delayed seroconversion^13,15^. Previously reported cohorts targeted patients with confirmed SARS-CoV-2 infection over a discrete period of time (both in terms of recruitment and antibody testing strategy)^10,11^. We may have been able to capture this with our longer follow up and denominator definition. Our longitudinal, rolling recruitment strategy aimed to cover a broad church of patients, and therefore may more accurately represent the cancer population as a whole. This will be an area of subsequent study as SCCAMP continues to evaluate the serological response of patients over time since vaccination.

We note some limitations of SCCAMP. Our study has not evaluated specific aspects of the immune response, including T-cell response or quantitative, longitudinal assessment of different antibody classes. This may reveal a differential in the longevity or robustness of the immune response to SARS-CoV-2 in patients treated with SACT. As regular asymptomatic PCR screening was not routine clinical practice during the studied period of time, we acknowledge that asymptomatic cases, particularly in the post-vaccination time period, will likely be underestimated. However, broadly we can still presume that our results are reflective of symptomatic infection, and our comments regarding COVID-19 PCR test results should be interpreted accordingly. In addition to this, as noted where relevant in this analysis, the number of patients with a positive SARS-CoV-2 PCR were relatively small, and consequently have been cautious in over-analysing sub-categories of this group in our cohort. Our real-world follow-up strategy inevitably results in not all patients providing all or as many samples for antibody testing. This is also reflective of the need to balance between exposing patients to contact only when needed and the dynamic research changes demanded by the waves of SARS-CoV-2 infection. Our study relies on publicly available and published data to provide control data for a non-cancer population, and although patients on other treatments not thought to significantly impact on the immune system (‘other’) have acted as a control group for our cohort, we acknowledge that some treatments in this category such as targeted therapies can impact on the immune system. Subsequent analysis on this cohort will include the trend of COVID-19 antibody over time, the effect of ACT and look at specific subgroups to explore the immune profile in greater detail.

## Conclusion

This preliminary report from SCCAMP suggests that in patients with cancer receiving ACT during the COVID-19 pandemic, symptomatic SARS-CoV-2 infection rates have been comparable to the general population. Significant protection is offered by vaccination, both in terms of antibody response and survival, and irrespective of type of ACT received. Vaccination against COVID-19 should be widely encouraged in patients with cancer undergoing treatment.

## Data Availability

All data produced in the present work are contained in the manuscript

## Author contributions

KP and PSH conceived of the study, designed and wrote the protocol, coordinated the study, conducted data interpretation and wrote the manuscript. JPH conducted data acquisition, data management, data interpretation and wrote the manuscript. MV led the data collection, data acquisition and coordinated all aspects of data and trial management. LA, PH, HMcV, LP and NW were involved with patient recruitment, sample acquisition and sample management. MB provided data analysis and acquisition support. HMcC was involved in all aspects of data analysis and interpretation. DAC and JDF provided guidance and support in the study design. EF conducted all SARS-Cov-2 antibody testing and was involved in planning and interpretation of the results. KT provided support and guidance in SARS-Cov-2 testing and analysis. PM designed and managed the sample management system. IB and MH contributed to the literature review and analysis. All the authors gave final approval for publication.

## Acknowledgements

This study was supported by grants from the Edinburgh Cancer Centre Research Fund (S07720) and Edinburgh and Lothians Health Foundation (EHLF). We thank the SCCAMP Advisory Group for their general guidance and advice (Appendix 1). JPH is funded by the CRUK Edinburgh Centre. We thank the patients who agreed to participate in this study.

## Appendix 1

**SCCAMP Advisory Group**

In addition to listed authors:

- Mark Baxter, Russell Petty - Dundee Cancer Centre, Ninewells Hospital and Dundee Medical School, Dundee, DD1 9SY, UK
- Paul Brennan - Centre for Regenerative Medicine and Cancer Research UK Edinburgh Centre, Institute for Regeneration and Repair, The University of Edinburgh, Edinburgh BioQuarter, 5 Little France Drive, Edinburgh EH16 4UU, UK
- Ruth Fullerton, Duncan McLaren, Iain Phillips, Paul Ramage, Stefan Symeonides - Edinburgh Cancer Centre, NHS Lothian, Crewe Road South, Edinburgh EH4 2XU, UK
- Margot Watson – Patient Representative Group

## Supplemental data

## Supplemental methods

### Full Study Design

The SCCAMP study protocol is available on https://cancer-data.ecrc.ed.ac.uk/projects/sccamp/sccamp-information-for-professionals/. Patients were eligible if they were over the age of 18 with a confirmed diagnosis of solid organ cancer, defined as cancer or metastasis in situ, and/or receiving cancer treatment (surgery, radiotherapy, hormone therapy, chemotherapy, targeted therapy, immunotherapy) in the last 12 months, and attending for outpatient Cancer Centre Care. Patients were not eligible if they had a concurrent haematological malignancy due to the different clinical profile of this cohort.

Patients consented to a Biobank (NHS Lothian NRS BioResource, BioBank SR1418, NHS Research Ethics Committee (REC):20/ES/0061) or the SCCAMP study (REC:20/ES/0076) when attending for anti-cancer treatment (ACT), primarily SACT, at the Edinburgh Cancer Centre (ECC) either at the Western General Hospital (WGH), Edinburgh or St John’s Hospital (SJH), Livingston, providing blood samples and consenting to anonymised review of routine clinical data.

Patients provided further blood samples up to a maximum of five over 1 year from consent (approx. +42 days, +84 days, +6 months, +1 year), when returning for further routine out-patient care. Patients were recruited throughout the period (Figure 1B) alongside follow-up sample acquisition. Although the protocol permitted patients to be recalled to provide samples as study visits, we prioritised fitting in samples with routine out-patient care to minimise additional visits for patients and consequently potential contact which might expose them to SARS-Cov-2 infection. Serum samples were tested via the validated Siemens Total (IgG/M and IgA) SARS-Cov-2 antibody assay at Ninewells Hospital, NHS Tayside (17,18).

### Data collation – cancer type stratification

Cancer type at recruitment was extracted and stratified into one of 8 groups based on the most dominant cancer types seen in the study as follows: breast, lung and chest, gynae, lower gastrointestinal (GI), upper GI, urological, Skin or other. “Other” cancer types include: head and neck (n=16), soft tissue sarcoma (n=8), cancer of unknown primary (n=8), prostate (n=5), cancer of the central nervous system (n=3) & neuroendocrine (n=1).

### Age adjustment of COVID-19 data

COVID-19 positive cases were defined as cases with a supporting positive PCR test. Publically available population COVID-19 data was accessed from Public Health Scotland data sources at [link] and monthly incident rates and cumulative total calculated for the combined local authorities in which the two hospital sites reside (the City of Edinburgh and West Lothian). As the ages of the patients in our cohort are all >25 years old, population COVID-19 data was age corrected to remove individuals under the age of 25. To do so, national data of daily COVID-19 infections, which is split by age groups 0-14, 15-19 and 20-24, were combined and compared to combined infections for age groups 25-44, 45-64 & 60+. Monthly COVID-19 infection ratios within individuals less than 25 years old and those over 25 years old were calculated and used as an adjustment factor for local population data.

### Stratification and analysis of COVID-19 antibody data

Patients were initially classified into 1 of 3 categories: i) no antibody (Ab) data available for patients (n=177; 23.1%), ii) Ab data only available prior to 14 days after 1st vaccination date (n= 242, 31.6%), iii) Ab data available >14d after 1st vaccination date (n=347, 45.3%). Patients in group iii were then further categorised to determine the antibody response to vaccination into

a. Ab response - if the first antibody sample collected after vaccination 1 was reactive,
b. Delayed Ab response - if a single (or more) antibody sample collected after vaccination 1 was non-reactive but a later Ab sample was positive in the absence of PCR confirmed COVID-19.
c. Non-reactive after partial vaccination - if all antibody samples collected after vaccination 1 were non-reactive but samples were not collected 14 days after 2nd vaccination.
d. Non-reactive after full vaccination - if all antibody samples collected after vaccination 1 were non-reactive and samples were collected >14 days after 2nd vaccination.

To calculate antibody responses in total and split by treatment type, we considered cases in groups a, b and d, and calculated percentages against these values.To calculate COVID-19 prevalence between fully and partial/non vaccinated states, we considered all cases with at least 1 available antibody test (unvaccinated group only) and/or a positive PCR test.

## Supplemental figures

**Figure S1.**
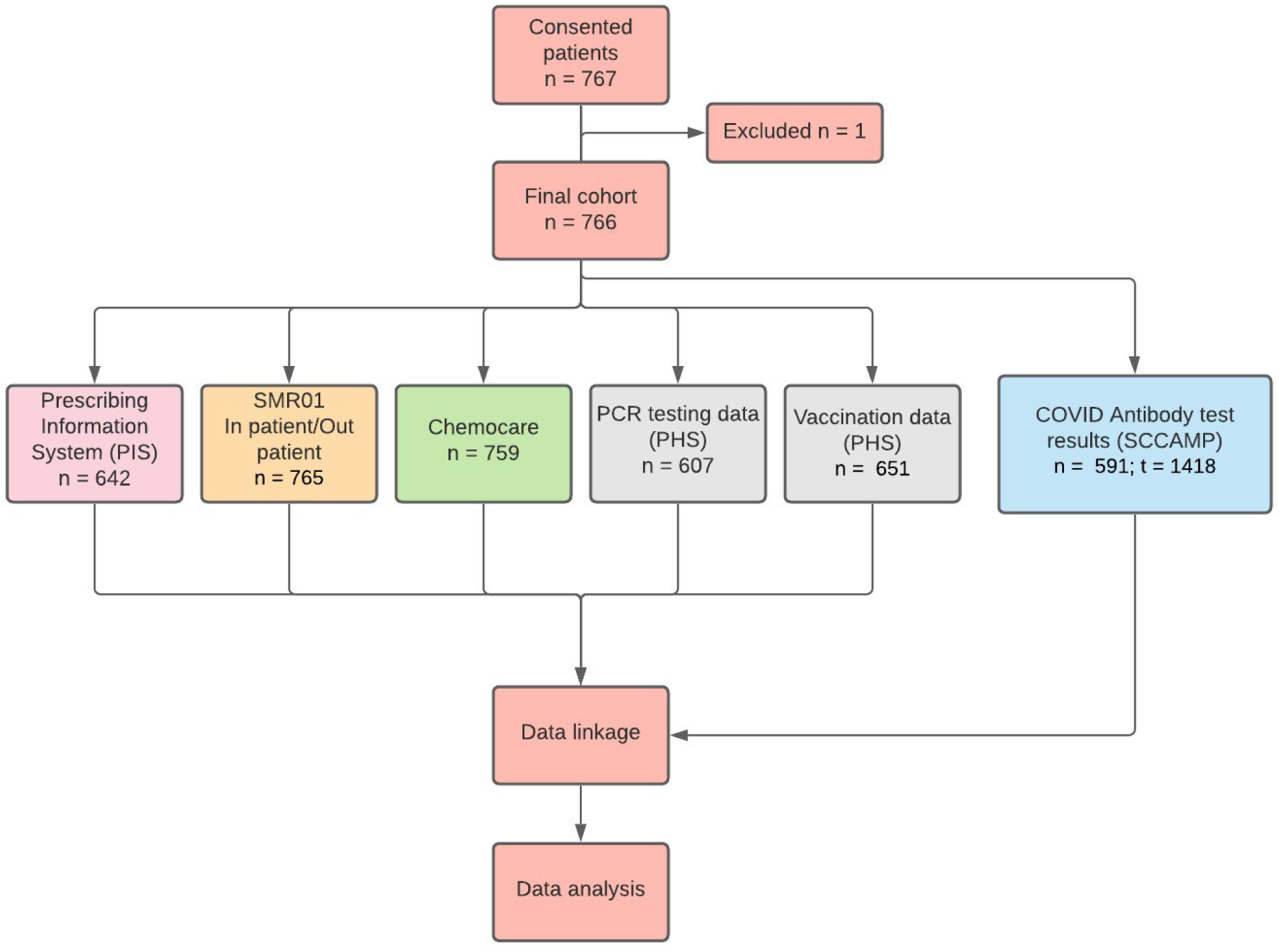
Summary of data linkage in SCCAMP. One patient was found to be diagnosed with concurrent haematological malignancy and excluded as per the exclusion criteria of the study. n = number of patients in each dataset. t = number of longitudinally collected antibody samples with processed data at time of data freeze.

**Figure S2.**
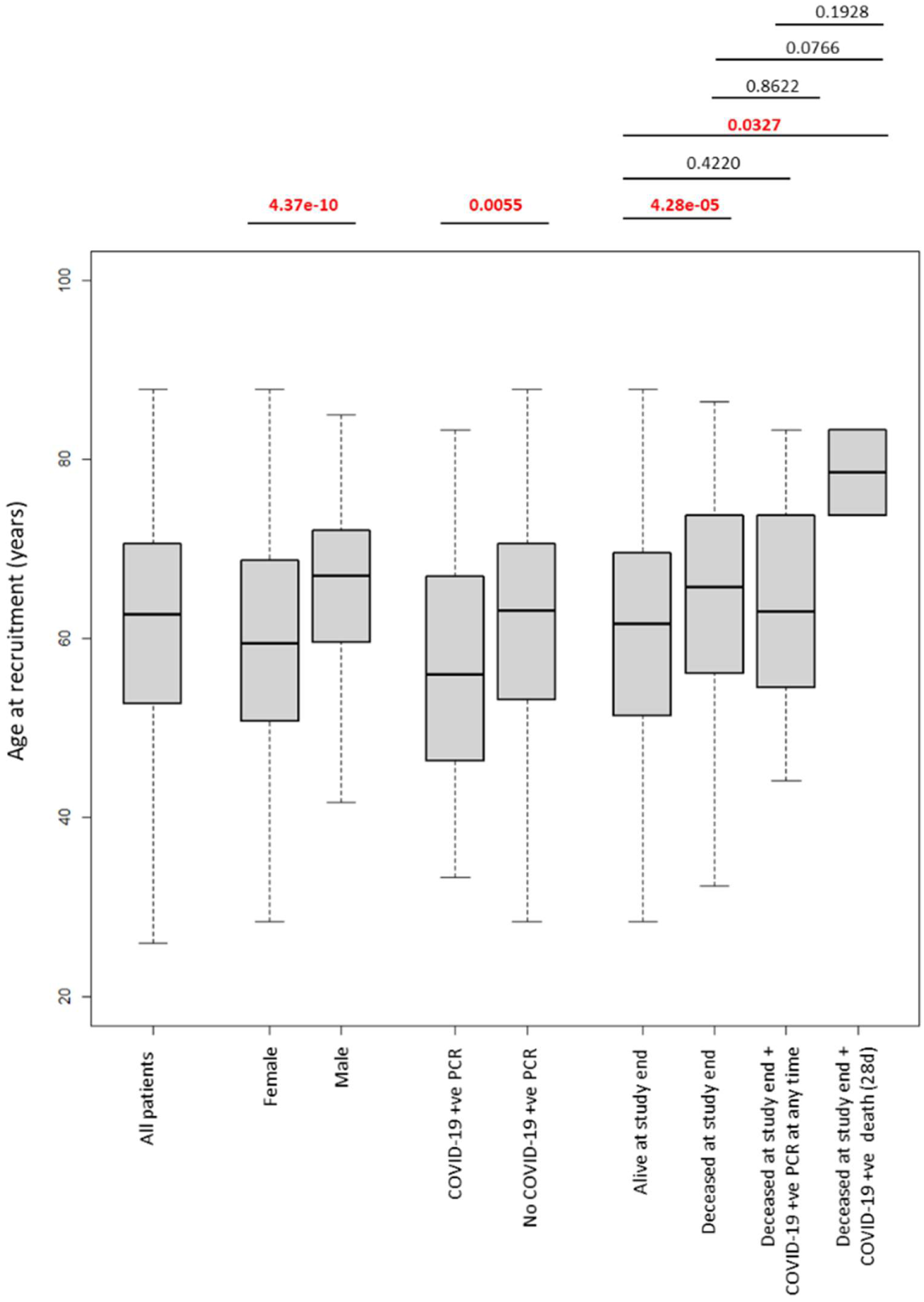
Boxplot of age of SCCAMP patients at recruitment split by a number of factors including gender, COVID-19 PCR status and survival outcome. P-values as calculated by pairwise Willcox tests are shown above with values in red reaching significance.

**Figure S3.**
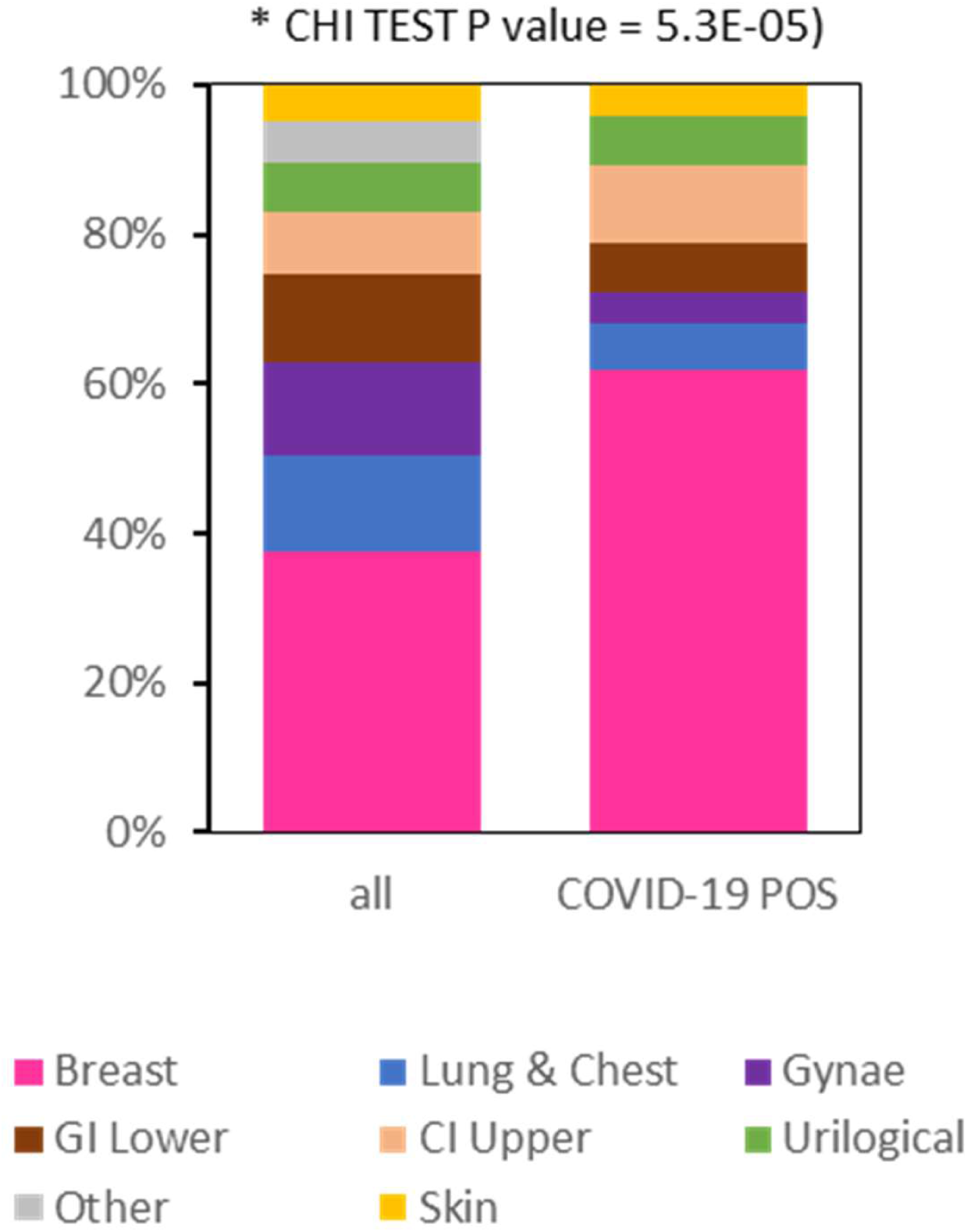
Stacked proportional bar plot of the cancer types present in the entire cohort (left) and those reported in patients with COVID-19 positive PCR. Chi squared test p-value comparing the relative proportion of cancer types between the two plots is shown above

**Figure S4.**
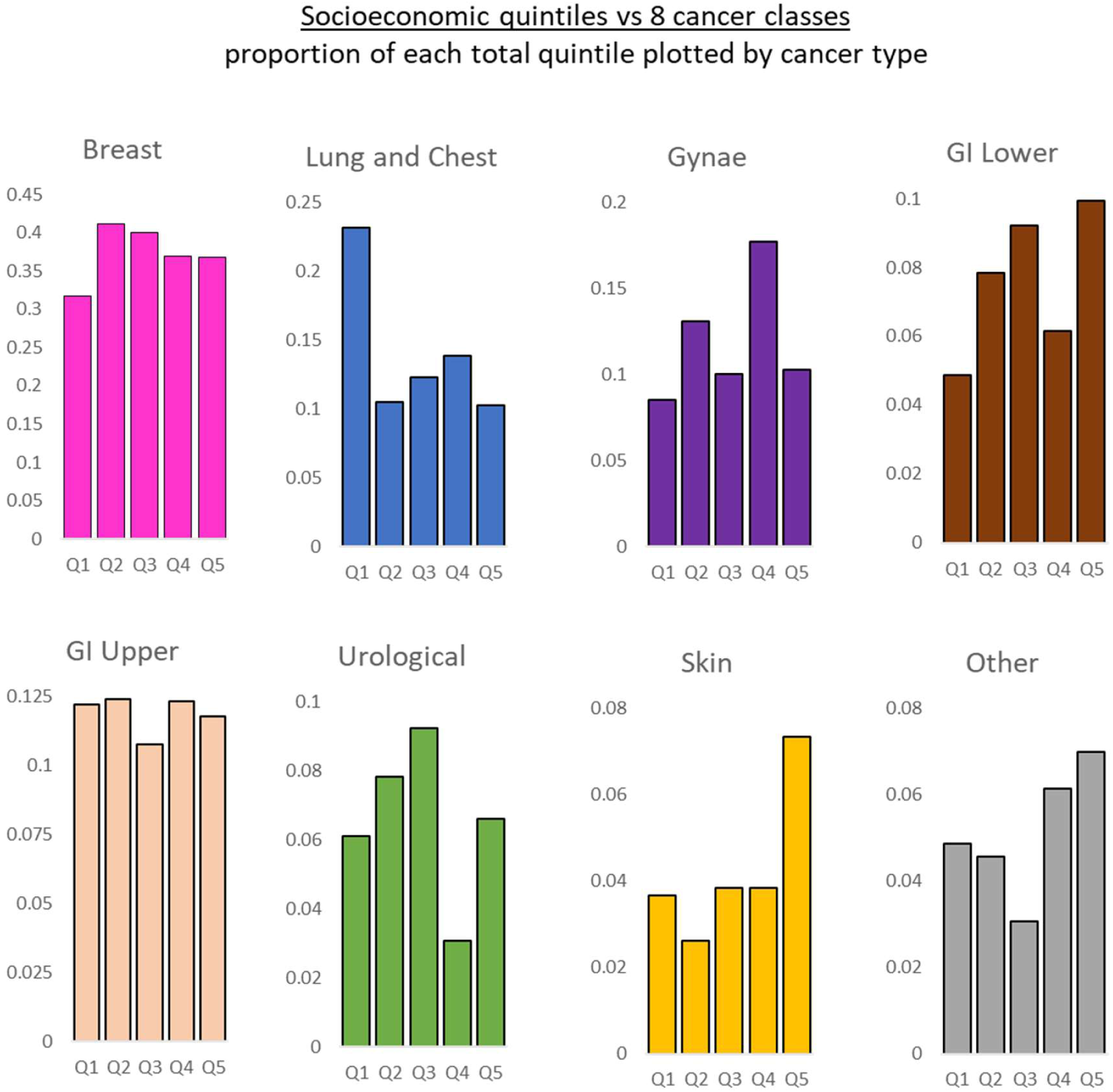
Plots of proportions total SMID socioeconomic quintiles found in each of the 8 cancer classifications.

**Figure S5.**
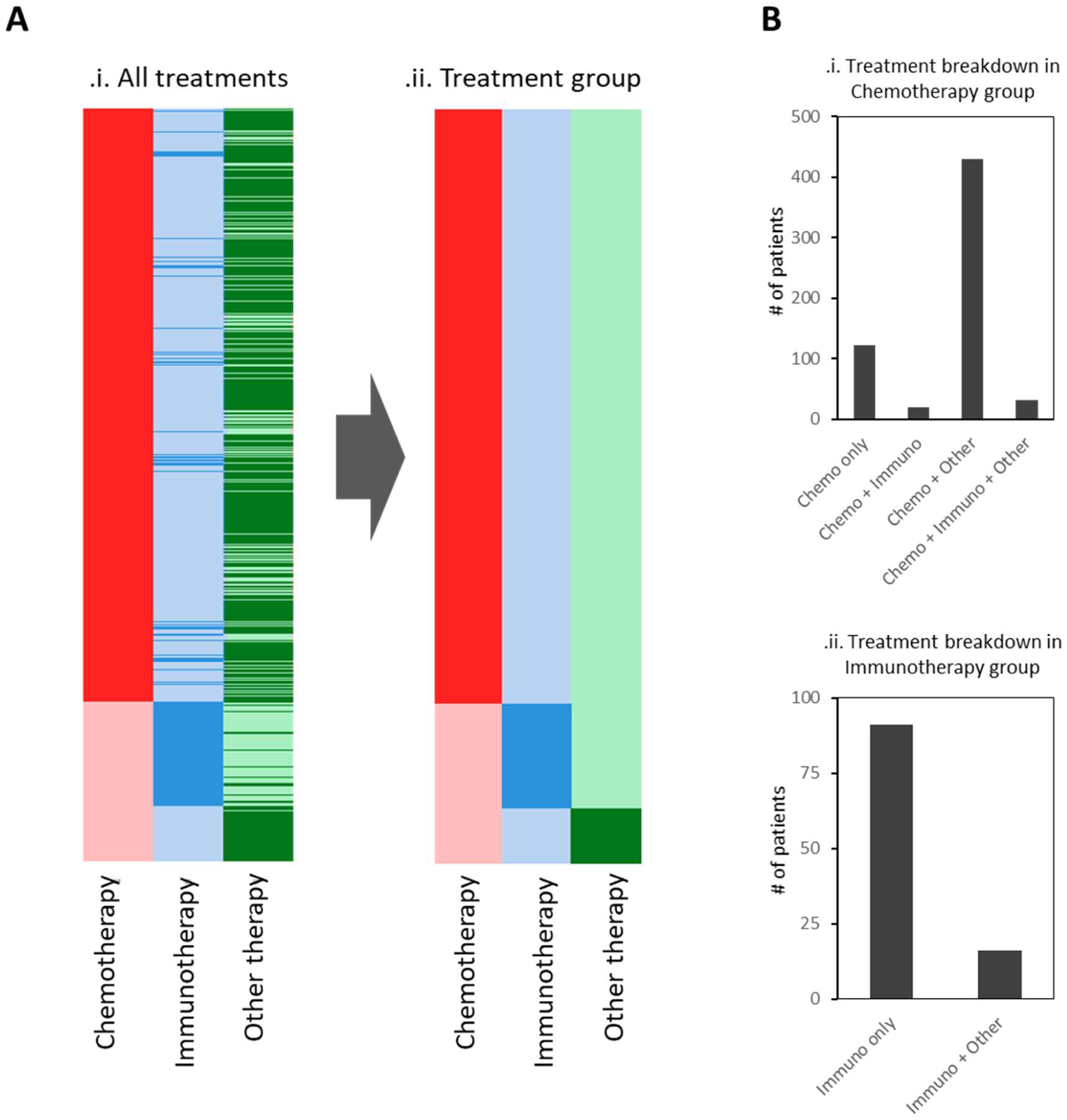
**A**. Heatmap plot of all treatment types across the 766 patients (i) as well as classified into the hierarchical treatment group (ii). **B**. Plots of treatment breakdown numbers for patients in the chemotherapy group (i) and immunotherapy group (ii).

**Figure S6.**
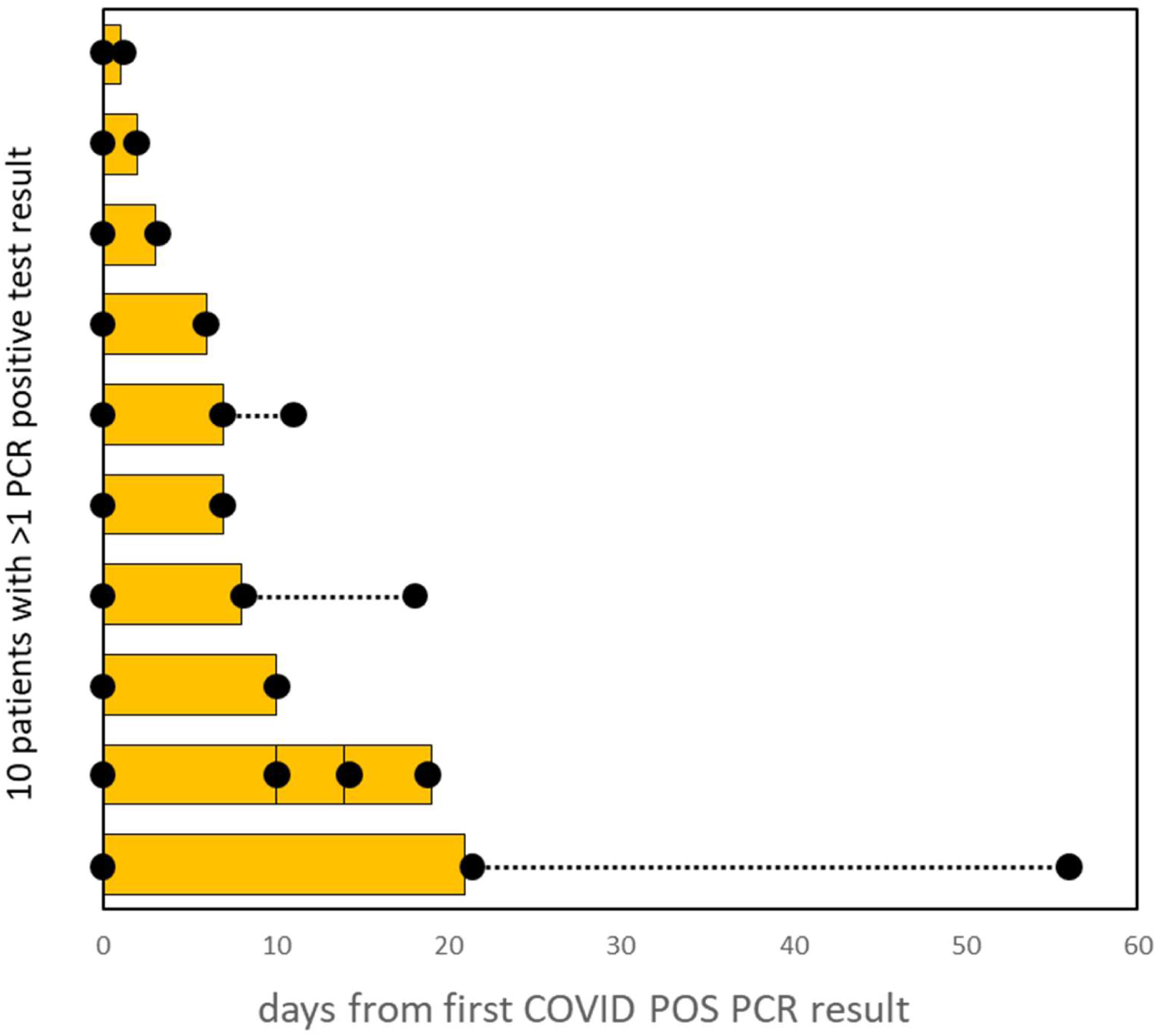
Plot of the number of days COVID-19 positive PCR result observed in patients with >1 positive PCR recorded. Black dots denote timing of PCR test. Dashed lines denote time between last positive test and first negative test.

**Figure S7.**
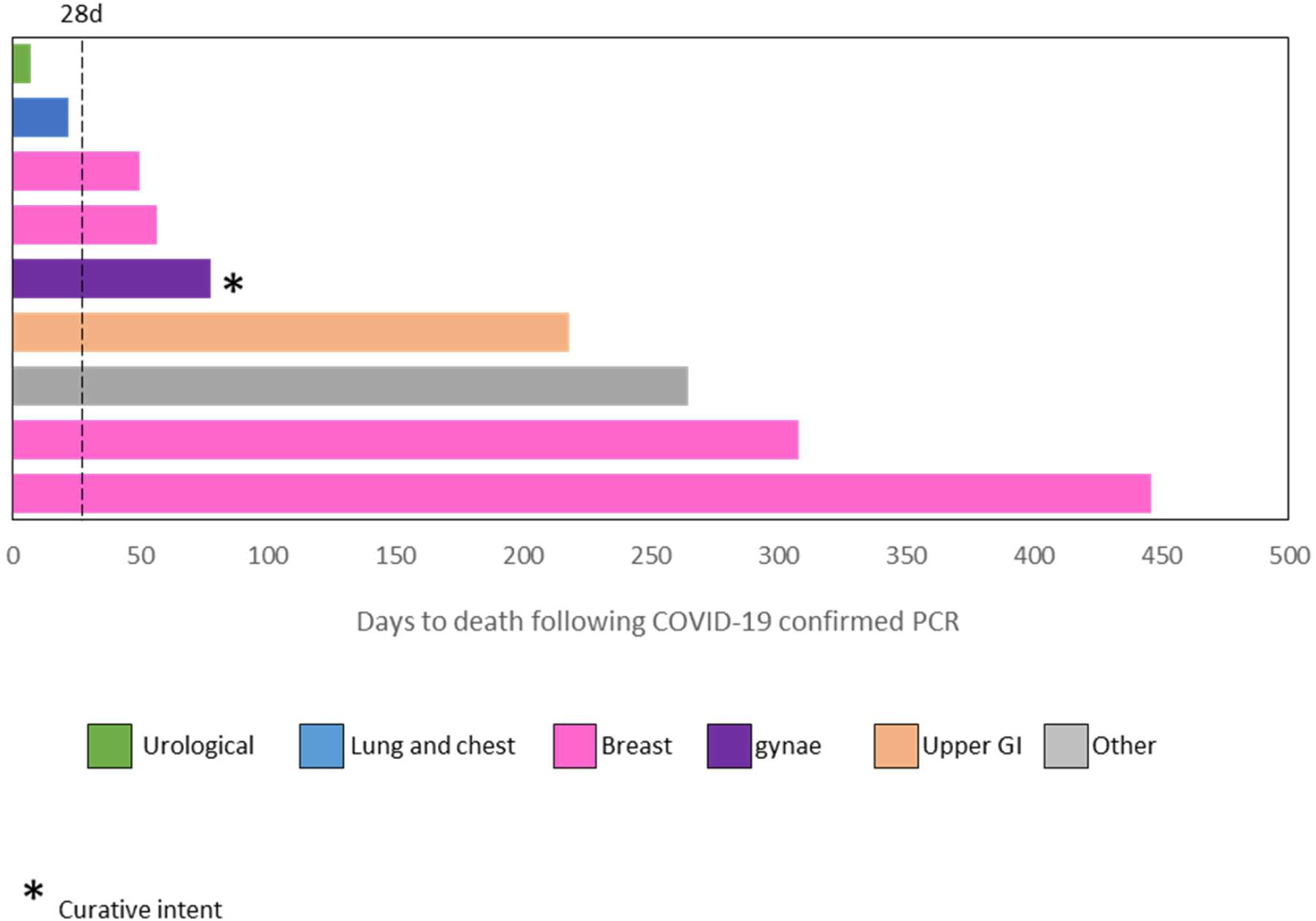
Plot of days to death in patients who were COVID-19 positive and then died during the study. Bars are colour coded by cancer type. Dashed bar denotes the classical classifier of a “COVID-19 death” set to 28 days after infection. Asterisk denotes curative intent.

**Figure S8.**
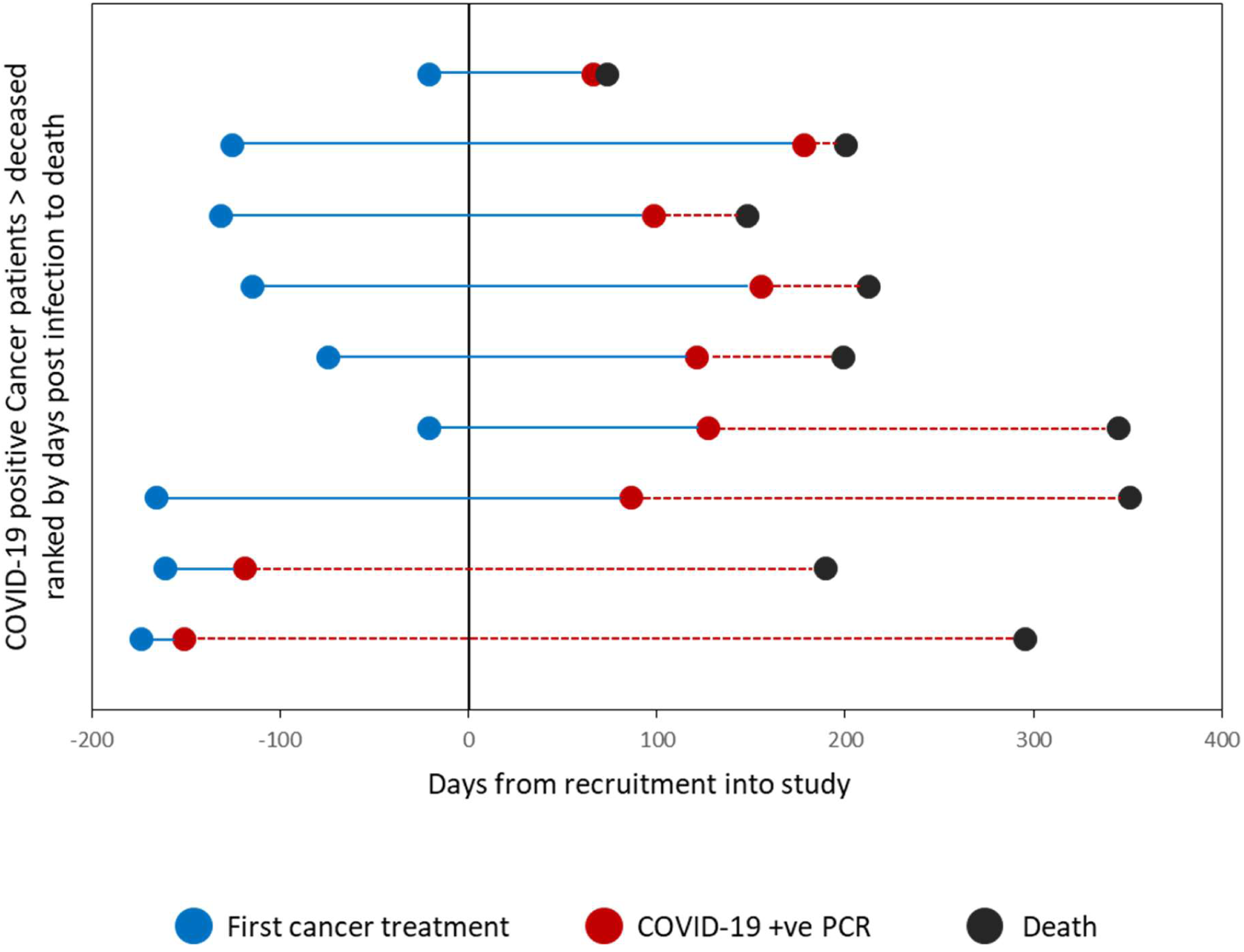
Plot of time between first cancer treatment (blue dots), COVID-19 infection (red dots) and death (black dots) across the 9 COVID-19 positive cancer patients in the study who went on to die before the study end. Plots are ranked by time between COVID-19 infection and death. Blue bars denote time between first treatment and infection, dashed red lines denote time between infection and death. Plots display time with respect to date of recruitment into the study.

**Figure S9.**
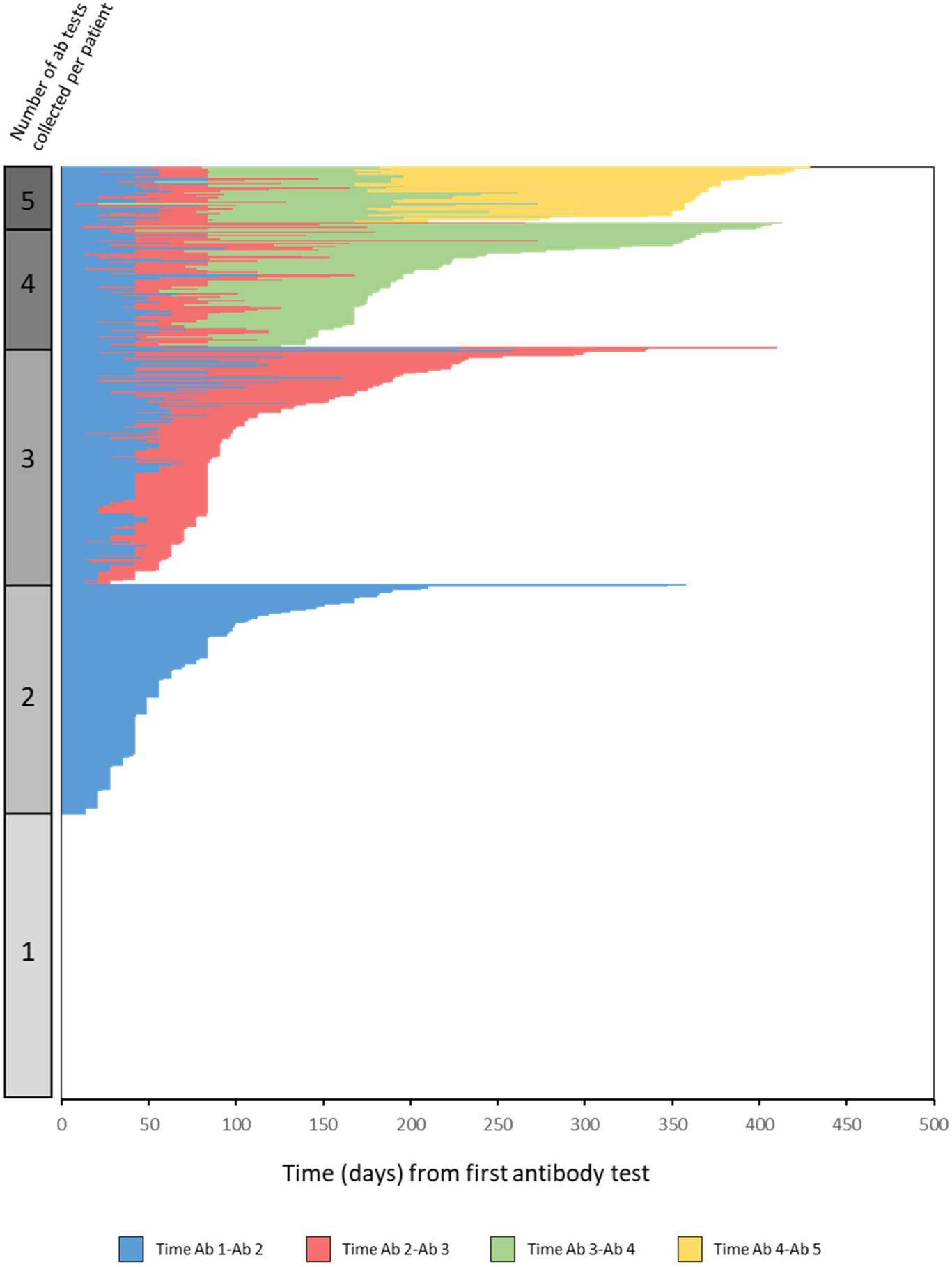
Plot of time differences between antibody test collections (days) for patients with available antibody data. Length of bars indicate time from previous collection. Note, no lines are visible for patients with only 1 antibody data point. Plot is ranked by length of time between antibody tests then ranked by number of collections.

**Figure S10.**
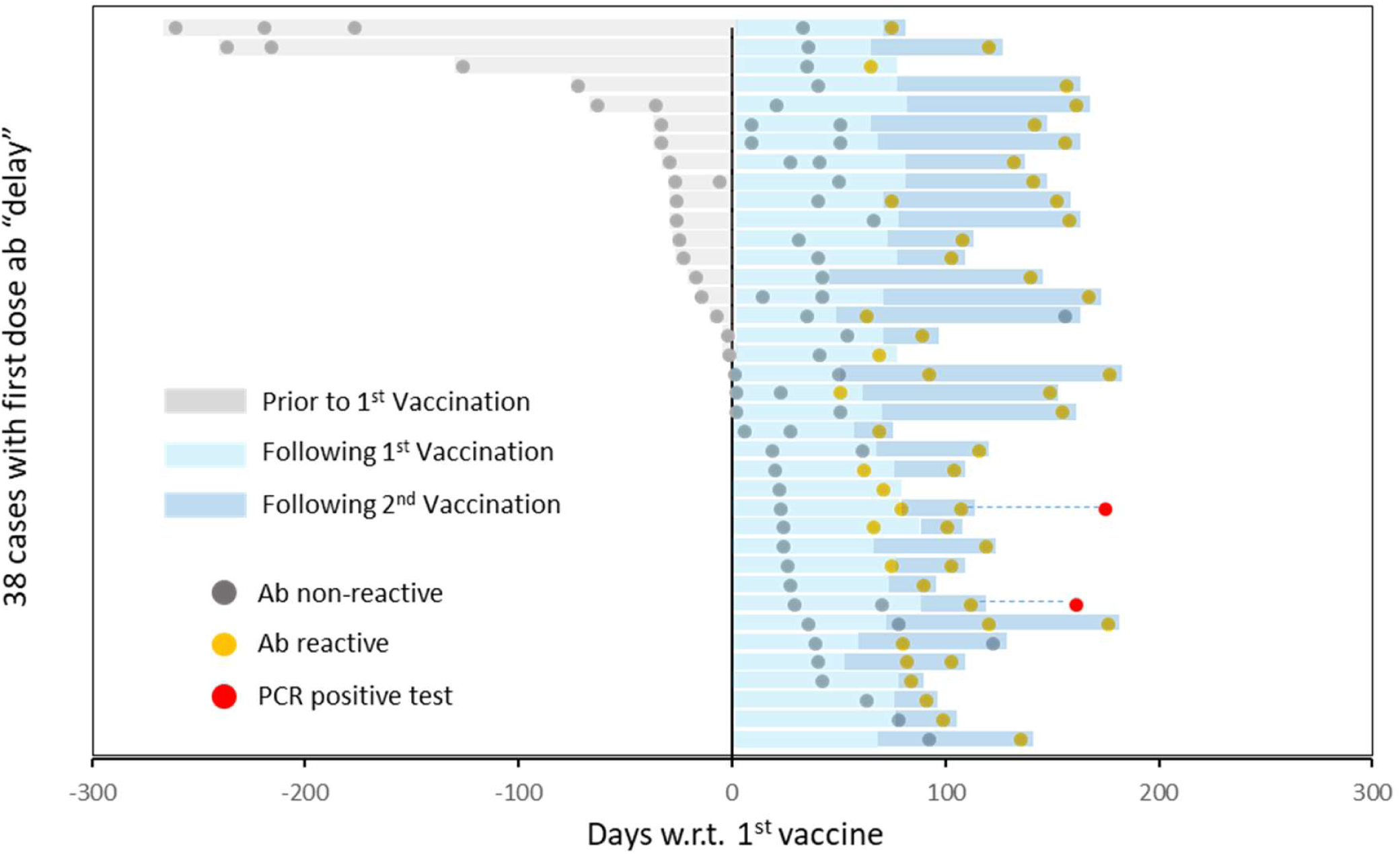
Plot of 38 cases displaying an initial non-reactive antibody result following vaccination 1, followed by a later conversion to reactive antibody state. All data is plotted with respect to (“w.r.t”) the date of vaccine 1. Dots denote antibody test collection dates. Grey dots = antibody non-reactive result, yellow dots = antibody reactive result, red dot = date of PCR positive test. Bars denote time w.r.t to vaccine dates. Grey bar = time prior to 1^st^ vaccination, light blue = time between 1^st^ vaccination and 2^nd^ vaccination (or last ab test), dark blue = time following 2^nd^ vaccination to last ab test. Dashed lines connecting to red dots denote time to PCR positive result. Cases are ranked by relative time to 1^st^ vaccine.

**Figure S11.**
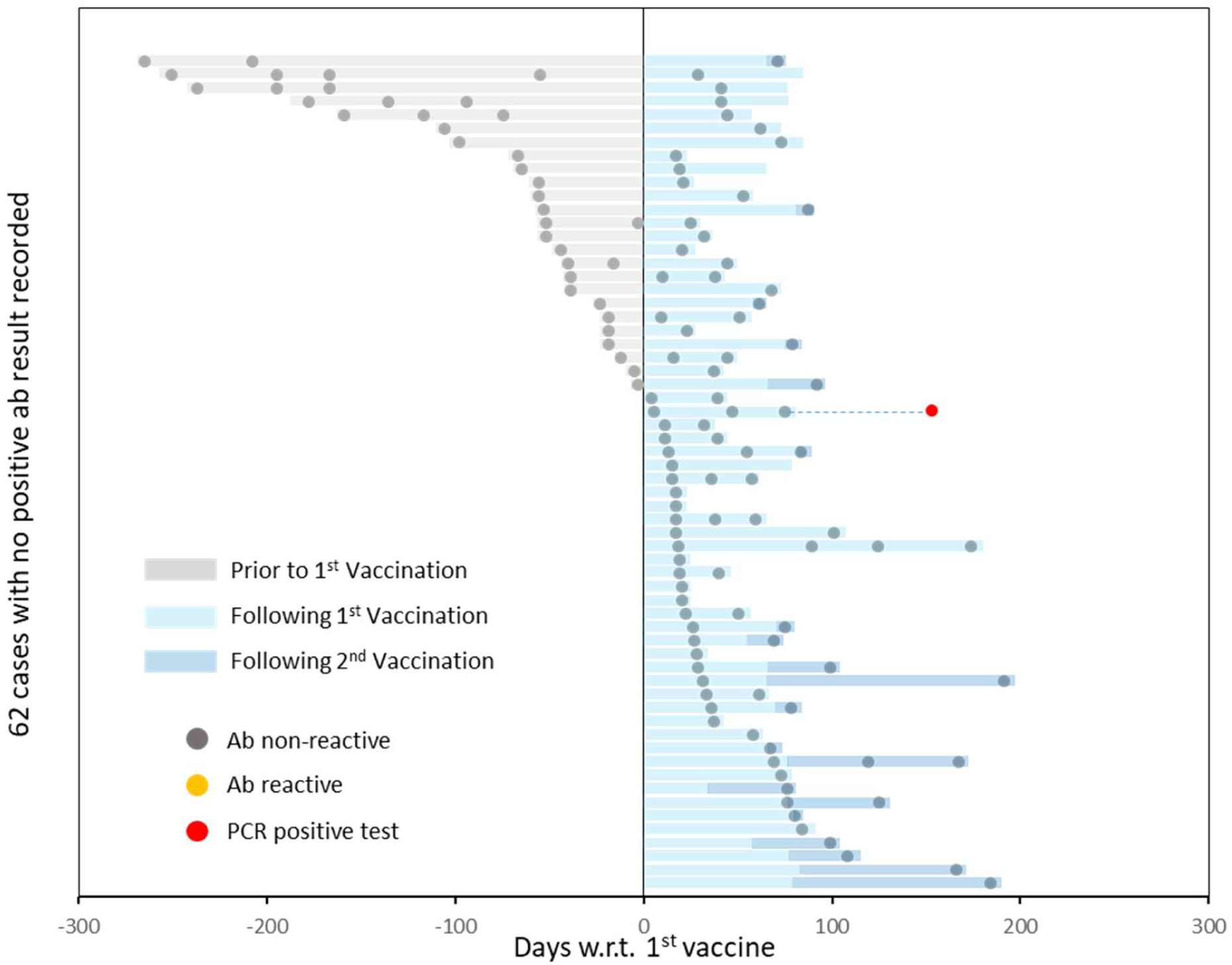
Plot of 62 cases displaying not reporting a reactive antibody result following vaccination 1. All data is plotted with respect to (“w.r.t”) the date of vaccine 1. Dots denote antibody test collection dates. Grey dots = antibody non-reactive result, yellow dots = antibody reactive result, red dot = date of PCR positive test. Bars denote time w.r.t to vaccine dates. Grey bar = time prior to 1^st^ vaccination, light blue = time between 1^st^ vaccination and 2^nd^ vaccination (or last ab test), dark blue = time following 2^nd^ vaccination to last ab test. Dashed lines connecting to red dots denote time to PCR positive result. Cases are ranked by relative time to 1^st^ vaccine.

**Figure S12.**
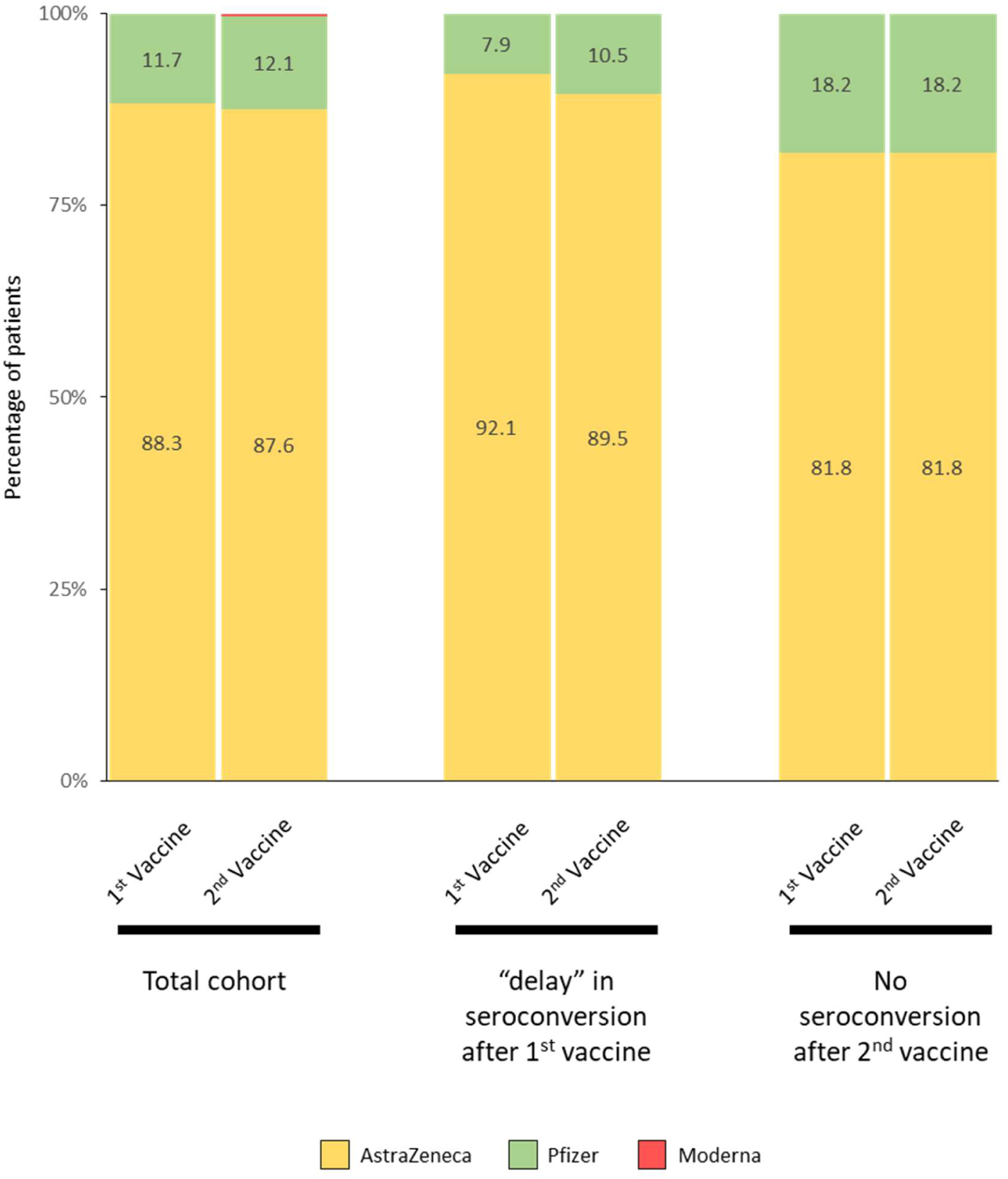
Stacked plots of percentage vaccine types split by manufacturer for the total cohort, patients displaying a “delayed” seroconversion following their first vaccination and patients displaying no seroconversion >14 days after their second vaccination.

**Figure S13.**
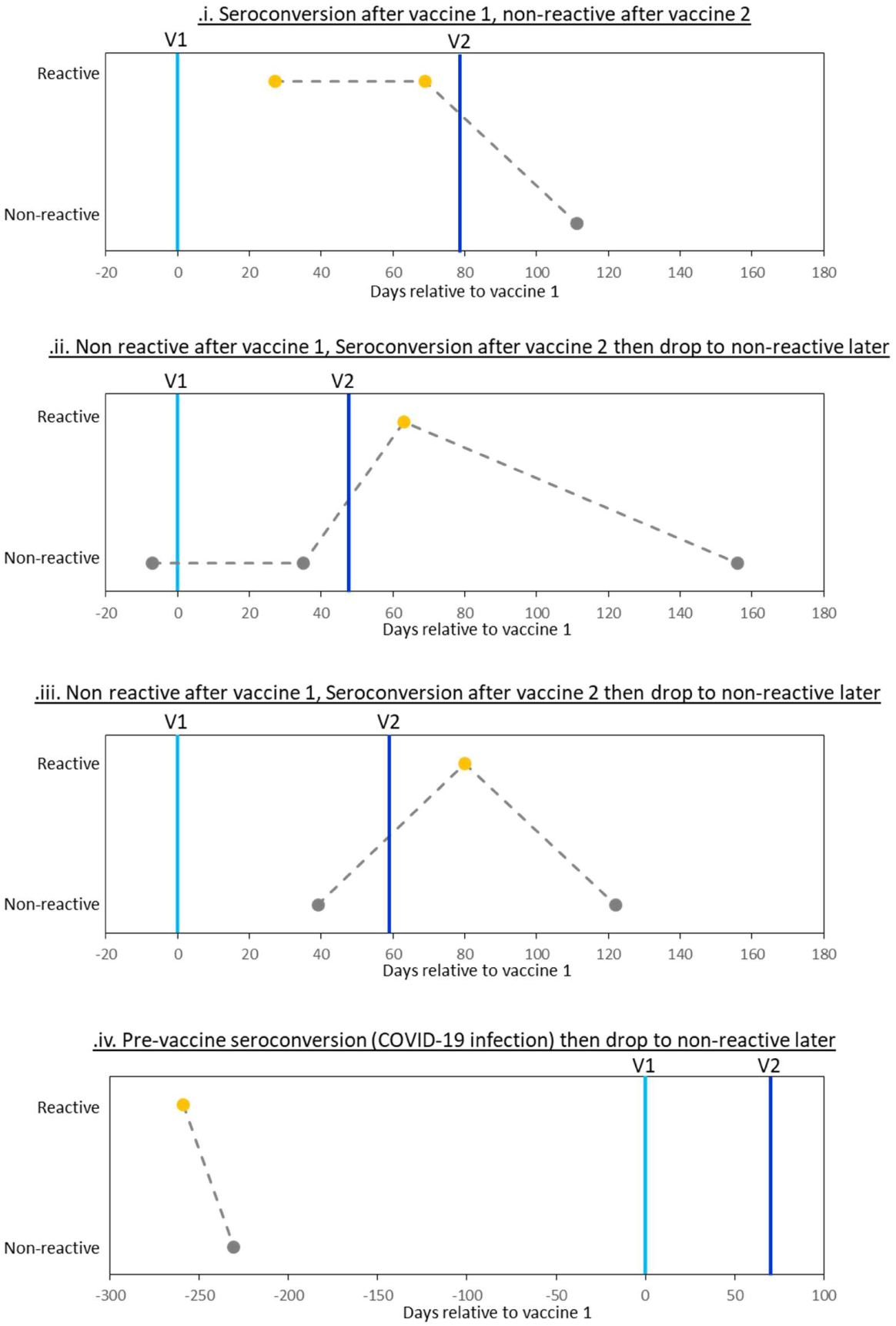
Plots of 4 cases displaying sero-reversion at any time point in the study. Cases i,ii and iii represent post vaccination sero-reversion. Plots denote binary results (“reactive” and “non-reactive”) plotted relative to the date of their first vaccination (light blue line; day = 0). Vaccination time point 2 is denoted by a dark blue line. Grey dots denote non-reactive results, yellow dots denote reactive results.

## Supplemental Tables

**Supplemental table 1.**
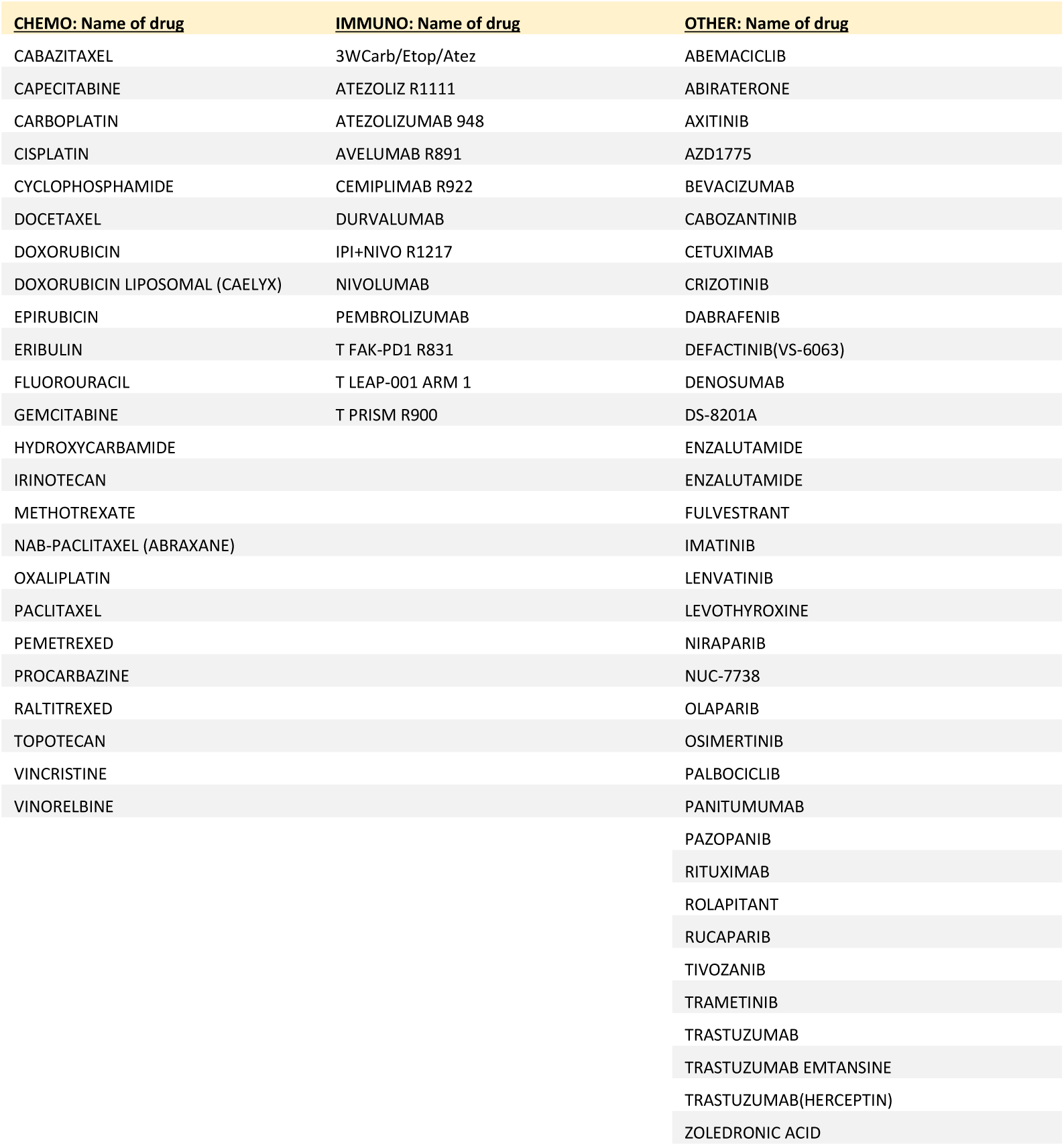
Table of drugs administered split by classification.

